# Artificial Intelligence-Driven Identification of Age- and Treatment-Specific TP53 and PI3K Alterations in Pancreatic Ductal Adenocarcinoma

**DOI:** 10.64898/2026.04.07.26350355

**Authors:** Fernando C. Diaz, Brigette Waldrup, Francisco G. Carranza, Sophia Manjarrez, Enrique Velazquez-Villarreal

**Affiliations:** Lineberger Comprehensive Cancer Center, University of North Carolina, Chapel Hill, NC, United States; City of Hope, Beckman Research Institute, Department of Integrative Translational Sciences, Duarte, CA; City of Hope Comprehensive Cancer Center, Duarte, CA

## Abstract

**Background:** Despite extensive characterization of key oncogenic drivers, pancreatic ductal adenocarcinoma (PDAC) continues to exhibit profound molecular heterogeneity and inconsistent responses to standard therapies, including gemcitabine. The role of pathway-level alterations, particularly in the context of age at onset and therapeutic exposure, remains insufficiently defined.

**Methods:** In this study, we leveraged a conversational artificial intelligence framework (AI-HOPE-TP53 and AI-HOPE-PI3K) to enable precision oncology, driven interrogation of clinical and genomic data from 184 PDAC tumors, stratified by age at diagnosis and gemcitabine exposure. Using AI-enabled cohort construction and pathway-centric analyses, we evaluated alterations in TP53 and PI3K signaling networks, with findings validated through conventional statistical methods.

**Results:** TP53 pathway analysis revealed a significantly higher frequency of TP53 mutations in early-onset compared to late-onset PDAC among gemcitabine-treated patients (86.7% vs. 57.1%, p = 0.04), with a similar trend observed between treated and untreated early-onset cases (86.7% vs. 40%, p = 0.07). Notably, in late-onset PDAC patients not treated with gemcitabine, absence of TP53 pathway alterations was associated with improved overall survival (p = 0.011). Complementary analyses of the PI3K pathway demonstrated a higher prevalence of pathway alterations in late-onset gemcitabine-treated tumors compared to untreated counterparts (13.2% vs. 2.7%, p = 0.02). Importantly, among late-onset patients not receiving gemcitabine, those without PI3K pathway alterations exhibited significantly improved overall survival (p < 0.0001).

**Conclusion:** Together, these findings identify distinct TP53 and PI3K pathway dependencies that are modulated by both age-of-onset and treatment exposure in PDAC. This work highlights the utility of conversational artificial intelligence in enabling rapid, integrative, and hypothesis-generating analyses within a precision oncology framework, supporting the identification of clinically relevant molecular stratification strategies for this aggressive disease.

## 1. Introduction

Late-stage diagnosis, profound molecular heterogeneity, and a highly immunosuppressive tumor microenvironment define pancreatic ductal adenocarcinoma (PDAC), contributing to its status as one of the most aggressive and therapeutically refractory malignancies (1–3). While gemcitabine-based regimens remain central to clinical management, their efficacy is limited by rapid onset of chemoresistance and context-dependent variability in outcomes (4–6). This resistance is mediated not only by tumor-intrinsic genetic alterations but also by intricate tumor–stroma interactions, including those involving cancer-associated fibroblasts (CAFs), immune cells, and extracellular matrix remodeling, which collectively regulate tumor evolution and treatment response (7–9).

Among the key molecular determinants of PDAC biology, alterations in tumor suppressor pathways and survival signaling networks are particularly critical. Mutations in TP53 occur in the majority of PDAC cases and are associated with genomic instability, impaired apoptosis, and enhanced tumor progression (1,10). Beyond its canonical tumor suppressor role, TP53 dysfunction has been increasingly linked to immune evasion mechanisms and remodeling of the tumor microenvironment, further contributing to poor clinical outcomes (11). In parallel, aberrant activation of the PI3K/AKT signaling pathway plays a central role in promoting tumor cell survival, metabolic adaptation, and resistance to cytotoxic therapies, including gemcitabine (4,12,13). Activation of this pathway has been shown to regulate key processes such as glycolysis, epithelial, mesenchymal transition (EMT), and drug efflux, thereby sustaining tumor aggressiveness and therapeutic escape (14,15).

Importantly, PI3K pathway dysregulation is not a static feature but evolves dynamically under therapeutic pressure. Multiple studies have demonstrated that inhibition of PI3K/AKT signaling can enhance gemcitabine sensitivity, while its activation contributes to resistance through mechanisms including metabolic reprogramming, anti-apoptotic signaling, and modulation of drug transport systems (4,16,17,18). Similarly, emerging evidence suggests that TP53 status may influence treatment response and disease progression in a context-dependent manner, highlighting the need to move beyond single-gene paradigms toward integrative, pathway-level analyses that reflect the complexity of PDAC biology (10,11).

These molecular processes are further modulated by the tumor microenvironment, where cytokine signaling and stromal interactions reinforce resistance phenotypes. For example, TGF-β-mediated signaling contributes to fibroblast activation, immune suppression, and chemoresistance, while cross-talk with pathways such as JAK/STAT and PI3K amplifies tumor-promoting signals (7,8,19). Additionally, chemotherapy itself can induce adaptive responses, including immune checkpoint upregulation and metabolic shifts, which may paradoxically support tumor persistence despite initial cytotoxic effects (20,21,22). Collectively, these findings underscore the necessity of understanding how key oncogenic and tumor suppressor pathways interact within clinically relevant contexts, including patient age and treatment exposure.

A critical challenge in precision oncology is the ability to systematically interrogate these complex interactions across large, heterogeneous clinical-genomic datasets (23–30). Traditional analytical frameworks often lack the flexibility to rapidly construct and refine multi-dimensional cohorts stratified by clinically meaningful variables such as age of onset and therapeutic exposure. This limitation is particularly relevant in PDAC, where early-onset and late-onset disease may represent biologically distinct entities with differential pathway dependencies and clinical trajectories (31,32).

We applied a conversational artificial intelligence framework to address this gap by enabling dynamic cohort construction and pathway-focused analysis of TP53 and PI3K alterations in PDAC. Building on prior AI-driven pathway interrogation approaches, AI-HOPE-TP53 (33) and AI-HOPE-PI3K (34), this method allows rapid hypothesis generation and testing across clinically stratified groups. We evaluated the distribution and clinical relevance of these alterations across age- and treatment-defined subgroups and assessed their associations with survival outcomes. These analyses aimed to uncover context-specific molecular dependencies that could guide precision oncology strategies, including combination therapies to overcome chemoresistance.

## 2. Materials and Methods

### 2.1 Study Design, Data Integration, and Clinical Stratification

This study was designed as a retrospective, integrative analysis of PDAC aimed at characterizing TP53 and PI3K pathway alterations across clinically relevant strata. A total of 184 PDAC tumor samples were included, each linked to curated clinical annotations, including age at diagnosis, treatment exposure, and survival outcomes. Molecular data were derived from tumor-based next-generation sequencing platforms, harmonized across sources to ensure consistency in variant annotation and gene-level interpretation.

To preserve patient-level independence, only one tumor profile per individual was retained. In cases with multiple sequencing records, a predefined prioritization strategy was applied, favoring samples with the most comprehensive genomic coverage and those temporally aligned with treatment exposure when available.

Patients were stratified by age at diagnosis into early-onset and late-onset groups using a uniform predefined threshold, selected to capture biologically meaningful differences in disease behavior. Treatment exposure was defined based on documented systemic therapy records, with patients classified as gemcitabine-exposed if they received gemcitabine-containing regimens at any point in their clinical course, and non-exposed otherwise. When temporal information was available, treatment status was aligned with sequencing timepoints to ensure concordance between therapeutic exposure and molecular profiling.

### 2.2 Pathway Definition, Genomic Annotation, and Statistical Analysis

Biologically informed gene panels were curated to represent (i) TP53-associated tumor suppressor signaling and (ii) the PI3K/AKT/mTOR signaling axis. The TP53 panel included TP53 and functionally related genes involved in DNA damage response, apoptosis, and cell cycle regulation. The PI3K panel encompassed catalytic subunits, regulatory components, and downstream effectors implicated in cell survival, metabolism, and therapeutic resistance.

Somatic variants were extracted from NGS datasets and filtered to retain protein-altering events, including missense mutations, nonsense variants, frameshift insertions/deletions, splice-site alterations, and start-loss mutations. Synonymous variants and non-coding alterations without predicted functional impact were excluded. Pathway alteration status at the patient level was defined by the presence of at least one qualifying mutation within the respective gene set. Gene-level alteration frequencies were calculated within clinically stratified subgroups to enable comparative analyses.

The primary molecular objective was to evaluate differences in TP53 and PI3K pathway alteration frequencies across age- and treatment-defined subgroups. Comparisons of categorical variables were performed using Fisher’s exact test or chi-square tests, as appropriate, with statistical significance defined as a two-sided p-value < 0.05.

Overall survival (OS) was defined as the interval from diagnosis to death or last follow-up and was analyzed using the Kaplan-Meier method, with group differences assessed by the log-rank test. Subgroup-specific survival analyses were conducted to evaluate the prognostic impact of pathway alterations within clinically relevant contexts, including interactions between age and gemcitabine exposure. All analyses were performed using validated computational pipelines to ensure reproducibility and adherence to standard oncologic statistical practices.

### 2.3 Artificial Intelligence–Enabled Analytical Framework

To enable scalable interrogation of complex clinicogenomic relationships, we implemented a conversational artificial intelligence framework based on the AI-HOPE platform. This system supports natural language–driven cohort construction, allowing dynamic generation of patient subgroups defined by combinations of clinical variables (e.g., age, treatment exposure) and molecular features (e.g., TP53 and PI3K alteration status).

Specialized modules, including AI-HOPE-TP53 and AI-HOPE-PI3K, were used for pathway-centric analyses, including mutation frequency aggregation, subgroup comparisons, and prioritization of candidate associations. The platform translates user-defined queries into executable analytical workflows, enabling iterative and reproducible hypothesis testing.

To ensure analytical rigor, all AI-generated outputs, including cohort definitions, mutation frequencies, and survival analyses, were independently validated using conventional statistical pipelines. Concordance between AI-derived and traditional analyses was systematically assessed to confirm data integrity, reproducibility, and methodological transparency.

## 3. Results

### 3.1 Baseline Clinical Profile of the PDAC Cohort

As shown in Table 1, the study cohort comprised 184 patients with PDAC, each with comprehensive demographic, clinical, and molecular annotations. All molecular analyses were derived from primary tumor specimens, ensuring consistency and comparability across the dataset.

**Table 1.** Overview of Baseline Demographic, Clinical, and Genomic Characteristics of the Study Cohort.

| Category | Subgroup | n (%) |
| --- | --- | --- |
| Age at Onset & Treatment Status | Early-Onset (<50), Gemcitabine-Treated | 15 (8.2%) |
| | Late-Onset ($\geq 50$), Gemcitabine-Treated | 91 (49.5%) |
| | Early-Onset ($< 50$), Non-Gemcitabine-Treated | 5 (2.7%) |
| | Late-Onset ($\geq 50$), Non-Gemcitabine-Treated | 73 (39.7%) |
| Tumor Classification | Pancreatic Adenocarcinoma | 184 (100.0%) |
| Sex Distribution | Male | 101 (54.9%) |
|  | Female | 83 (45.1%) |
| Specimen Type | Primary Tumor | 184 (100.0%) |
| Stage at Diagnosis | Stage I | 21 (11.4%) |
|  | Stage II | 151 (82.1%) |
|  | Stage III | 5 (2.7%) |
|  | Stage IV | 5 (2.7%) |
|  | Not Available | 2 (1.1%) |
| Ethnicity | Hispanic or Latino | 5 (2.7%) |
|  | Not Hispanic or Latino | 136 (73.9%) |
|  | Unknown | 43 (23.4%) |

The cohort was predominantly composed of patients with late-onset PDAC (≥50 years), with gemcitabine-treated individuals representing the largest subgroup, followed by untreated cases. Early-onset PDAC (<50 years) constituted a smaller fraction of the cohort. This distribution reflects both the higher prevalence of late-onset disease and the central role of gemcitabine-based regimens in clinical practice.

Sex distribution was relatively balanced, with a slight predominance of male patients. All samples were obtained from primary tumors, minimizing potential confounding from metastatic heterogeneity or treatment-induced clonal evolution.

Disease stage at diagnosis was primarily stage II, with fewer cases in early-stage disease (stage I) and limited representation of advanced stages (stage III and IV). A small subset of patients had unavailable staging information.

Most patients were classified as not Hispanic or Latino, with a smaller proportion identified as Hispanic or Latino and a notable fraction with unknown ethnicity.

Together, these baseline characteristics define a clinically representative PDAC cohort and provide the foundation for subsequent analyses of age- and treatment-specific patterns of TP53 and PI3K pathway alterations and their association with clinical outcomes.

### 3.2 Patterns of TP53 and PI3K Pathway Alterations Across Age and Treatment Contexts

Stratified analysis by age and gemcitabine exposure (Table 2a–d) revealed divergent patterns between TP53 and PI3K pathways. TP53 alterations were pervasive across all subgroups, while PI3K alterations demonstrated pronounced context-dependent variability, most notably in late-onset PDAC.

**Table 2.**
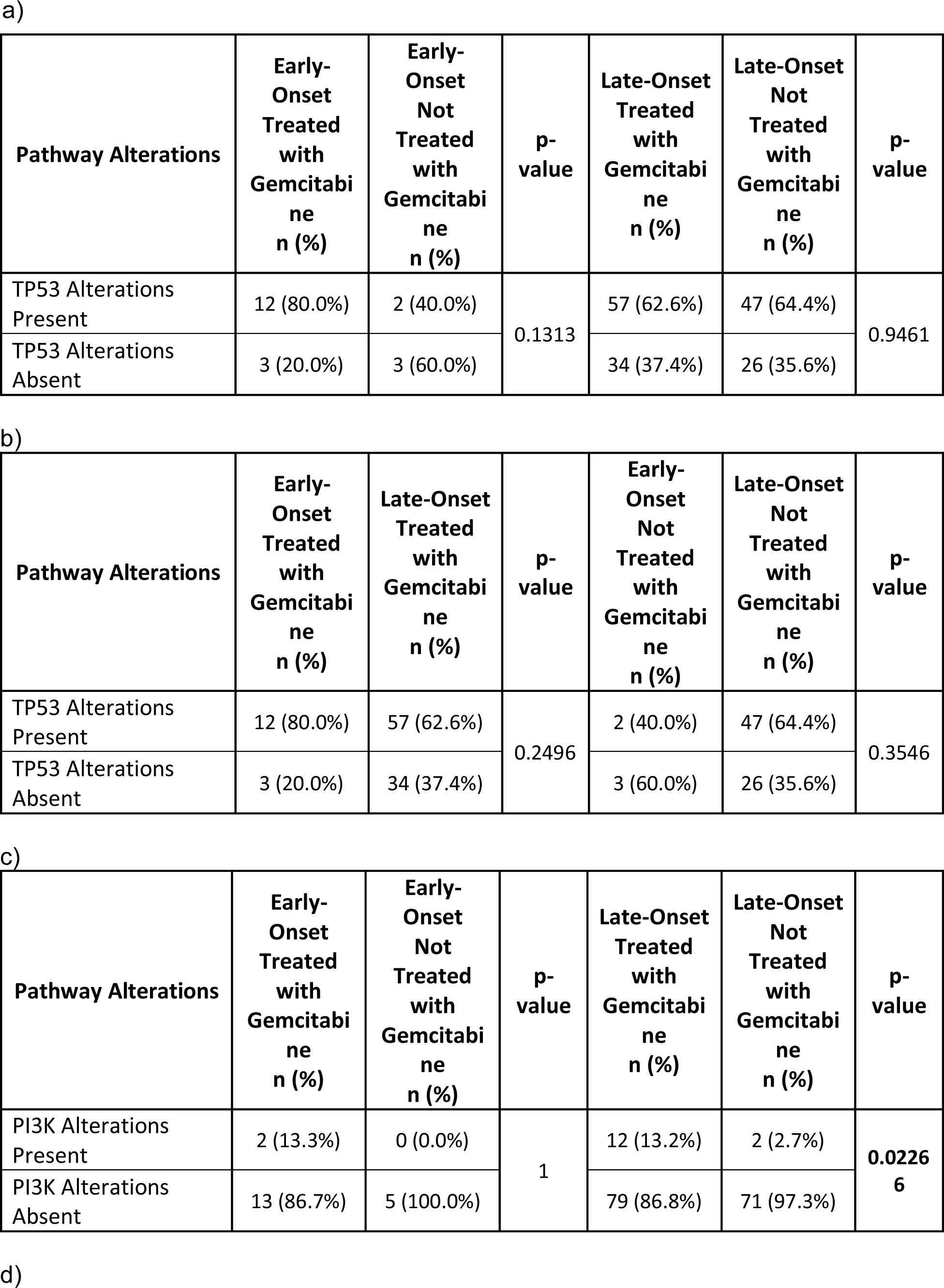
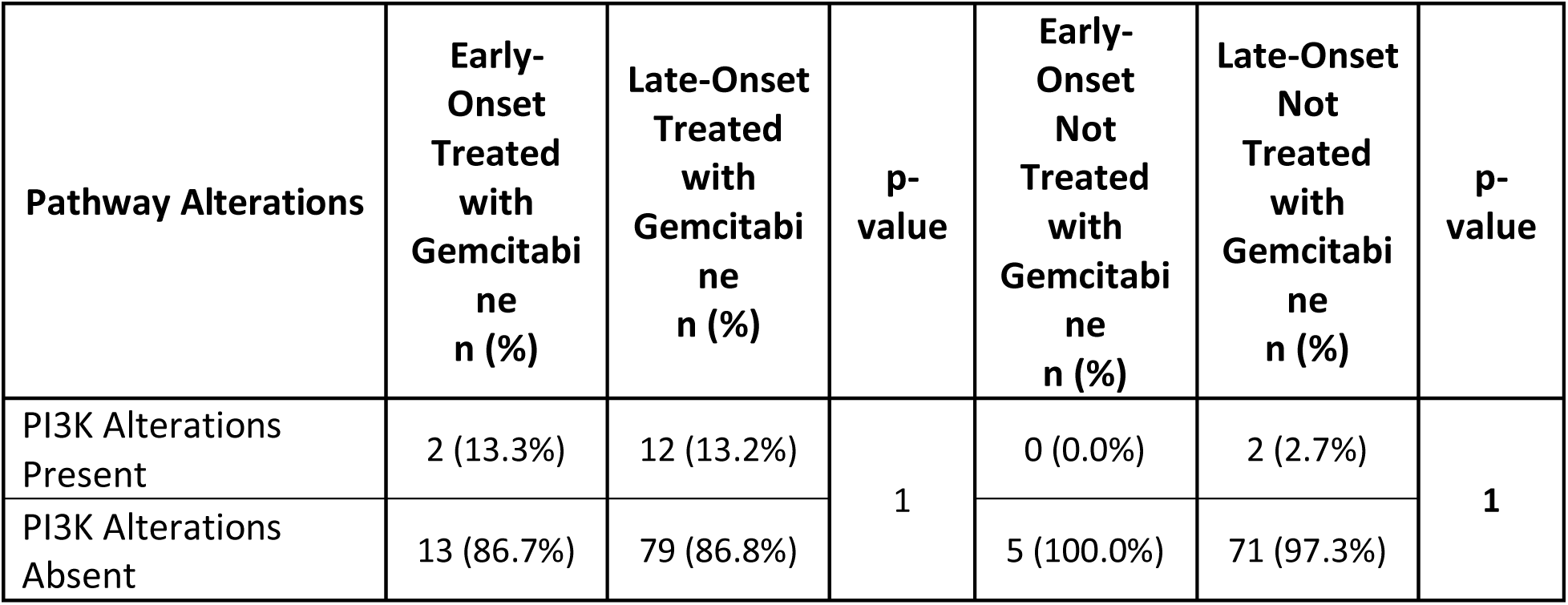
Distribution of TP53 and PI3K Pathway Alterations Across Age- and Treatment-Stratified PDAC Subgroups. This table summarizes the frequency of TP53 and PI3K pathway alterations across pancreatic ductal adenocarcinoma (PDAC) cohorts stratified by age at diagnosis (early-onset vs. late-onset) and gemcitabine exposure. The analysis is organized into four complementary panels to facilitate pathway-specific and clinically contextual interpretation: (a) TP53 pathway alterations comparing gemcitabine-treated and non-gemcitabine-treated tumors within early-onset PDAC; (b) TP53 pathway alterations comparing early-onset and late-onset disease within each treatment group; (c) PI3K pathway alterations comparing gemcitabine-treated and non-treated tumors within early-onset PDAC; and (d) PI3K pathway alterations comparing early- and late-onset PDAC within each treatment stratum.

| Pathway Alterations | Early-Onset<br>Treated<br>with<br>Gemcitabi<br>ne<br>n (%) | Early-Onset<br>Not<br>Treated<br>with<br>Gemcitabi<br>ne<br>n (%) | p-value | Late-Onset<br>Treated<br>with<br>Gemcitabi<br>ne<br>n (%) | Late-Onset<br>Not<br>Treated<br>with<br>Gemcitabi<br>ne<br>n (%) | p-value |
| --- | --- | --- | --- | --- | --- | --- |
| TP53 Alterations<br>Present | 12 (80.0%) | 2 (40.0%) | 0.1313 | 57 (62.6%) | 47 (64.4%) | 0.9461 |
| TP53 Alterations<br>Absent | 3 (20.0%) | 3 (60.0%) |  | 34 (37.4%) | 26 (35.6%) |  |

b)
| Pathway Alterations | Early-Onset<br>Treated<br>with<br>Gemcitabi<br>ne<br>n (%) | Late-Onset<br>Treated<br>with<br>Gemcitabi<br>ne<br>n (%) | p-value | Early-Onset<br>Not<br>Treated<br>with<br>Gemcitabi<br>ne<br>n (%) | Late-Onset<br>Not<br>Treated<br>with<br>Gemcitabi<br>ne<br>n (%) | p-value |
| --- | --- | --- | --- | --- | --- | --- |
| TP53 Alterations<br>Present | 12 (80.0%) | 57 (62.6%) | 0.2496 | 2 (40.0%) | 47 (64.4%) | 0.3546 |
| TP53 Alterations<br>Absent | 3 (20.0%) | 34 (37.4%) |  | 3 (60.0%) | 26 (35.6%) |  |

c)
| Pathway Alterations | Early-Onset<br>Treated<br>with<br>Gemcitabi<br>ne<br>n (%) | Early-Onset<br>Not<br>Treated<br>with<br>Gemcitabi<br>ne<br>n (%) | p-value | Late-Onset<br>Treated<br>with<br>Gemcitabi<br>ne<br>n (%) | Late-Onset<br>Not<br>Treated<br>with<br>Gemcitabi<br>ne<br>n (%) | p-value |
| --- | --- | --- | --- | --- | --- | --- |
| PI3K Alterations<br>Present | 2 (13.3%) | 0 (0.0%) | 1 | 12 (13.2%) | 2 (2.7%) | 0.0226<br>6 |
| PI3K Alterations<br>Absent | 13 (86.7%) | 5 (100.0%) |  | 79 (86.8%) | 71 (97.3%) |  |

d)
| Pathway Alterations | Early-Onset Treated with Gemcitabine n (%) | Late-Onset Treated with Gemcitabine n (%) | p-value | Early-Onset Not Treated with Gemcitabine n (%) | Late-Onset Not Treated with Gemcitabine n (%) | p-value |
| --- | --- | --- | --- | --- | --- | --- |
| PI3K Alterations Present | 2 (13.3%) | 12 (13.2%) | 1 | 0 (0.0%) | 2 (2.7%) | 1 |
| PI3K Alterations Absent | 13 (86.7%) | 79 (86.8%) |  | 5 (100.0%) | 71 (97.3%) |  |

#### 3.2.1 TP53 pathway alterations across age and treatment strata

TP53 pathway alterations were highly prevalent across the cohort, consistent with their central role in PDAC biology. Within early-onset PDAC, TP53 alterations were observed in 80.0% of gemcitabine-treated tumors compared to 40.0% in non-treated tumors, although this difference did not reach statistical significance (p=0.1313). In late-onset PDAC, TP53 alterations were similarly frequent in both treatment groups, occurring in 62.6% of gemcitabine-treated and 64.4% of non-treated tumors (p=0.9461) (Table 2a). Comparisons across age groups within treatment strata revealed no statistically significant differences. Among gemcitabine-treated patients, TP53 alterations were present in 80.0% of early-onset tumors and 62.6% of late-onset tumors (p=0.2496). In the non-treated setting, alteration frequencies were 40.0% in early-onset PDAC and 64.4% in late-onset PDAC (p=0.3546) (Table 2b). TP53 pathway disruption emerged as a pervasive feature of PDAC, remaining consistent across age groups and treatment contexts, with no statistically significant differences observed at the pathway level.

#### 3.2.2 PI3K pathway alterations across age and treatment strata

In contrast to TP53, PI3K pathway alterations were less frequent overall but demonstrated notable variability depending on clinical context. In early-onset PDAC, PI3K alterations were identified in 13.3% of gemcitabine-treated tumors and were absent in non-treated tumors (0.0%), although this difference was not statistically significant (p=1). In late-onset PDAC, a distinct pattern emerged. PI3K pathway alterations were detected in 13.2% of gemcitabine-treated tumors compared to only 2.7% of non-treated tumors, representing a statistically significant difference (p=0.02266) (Table 2c). This finding suggests a potential association between PI3K pathway activation and gemcitabine exposure in late-onset disease.

Comparisons of PI3K alteration frequencies across age groups within treatment strata did not reveal significant differences. Among gemcitabine-treated patients, alteration rates were nearly identical between early-onset and late-onset PDAC, while in the non-treated group, PI3K alterations were absent in early-onset tumors and detected only at low frequency in late-onset cases (Table 2d).

Together, these results indicate divergent pathway behavior in PDAC. While TP53 alterations are broadly consistent across subgroups, PI3K alterations appear more context-dependent, with enrichment in gemcitabine-treated late-onset disease, suggesting a role in treatment-related molecular adaptation.

### 3.3 Gene-Level landscape of TP53 and PI3K

Given the relative uniformity of TP53 pathway alterations and the context-dependent variability observed for PI3K signaling at the pathway level (Section 3.2), we next examined these pathways at single-gene resolution to uncover clinically relevant molecular patterns masked by aggregate analyses (Tables S1–S8). Across the cohort, TP53 emerged as the predominant alteration within the tumor suppressor axis, consistently detected across both early- and late-onset PDAC and irrespective of gemcitabine exposure, reinforcing its role as a foundational driver of genomic instability. In contrast, alterations within the PI3K pathway were less frequent but distributed across multiple genes, including catalytic and regulatory components, suggesting a more heterogeneous and context-dependent architecture of pathway activation. Importantly, discrete gene-level events within the PI3K signaling network appeared enriched in specific clinical subgroups, particularly in gemcitabine-exposed tumors, highlighting potential adaptive mechanisms linked to therapeutic pressure. These findings reveal that, while TP53 alterations define a ubiquitous backbone of PDAC biology, PI3K pathway dysregulation is shaped by both age and treatment context, uncovering molecular dependencies that are not evident at the pathway level alone.

#### 3.3.1 TP53 gene-level landscape reveals age-associated enrichment in gemcitabine-treated early-onset PDAC

At the single-gene level, the TP53 pathway was dominated by alterations in TP53, with other pathway components occurring at low frequency across all subgroups. In early-onset PDAC, TP53 mutations were more frequently observed in gemcitabine-treated tumors (86.7%) compared to non-treated tumors (40.0%), representing a notable, although not statistically significant, difference (p=0.0726) (Table S1). Other genes within the TP53 pathway, including MDM2, MDM4, CDKN1A, ATM, CHEK1/2, PTEN, and BBC3, were largely unaltered in this subgroup, with only sporadic events such as CDKN2A (13.3-20.0%) and ATR (6.7%) detected at low frequencies.

In contrast, late-onset PDAC demonstrated a more uniform distribution of TP53 mutations across treatment groups. TP53 alterations were present in 57.1% of gemcitabine-treated tumors and 60.3% of non-treated tumors (p=0.8064), indicating no treatment-associated enrichment (Table S2). Similarly, other TP53 pathway genes exhibited low mutation frequencies without meaningful differences between treated and untreated cases. Notably, CDKN2A mutations were observed in approximately one-fifth of late-onset tumors (∼19-20%), while alterations in genes such as ATM, ATR, and CHEK2 occurred infrequently.

When comparing age groups within the gemcitabine-treated cohort, a statistically significant difference emerged. TP53 mutations were significantly more prevalent in early-onset tumors (86.7%) compared to late-onset tumors (57.1%) (p=0.0431) (Table S3), suggesting an age-dependent enrichment of TP53 alterations in the context of gemcitabine exposure. In contrast, no significant age-based differences were observed in the non-treated setting, where TP53 mutation frequencies were 40.0% in early-onset and 60.3% in late-onset PDAC (p=0.396) (Table S4).

Overall, these findings indicate that while TP53 mutations represent a central and recurrent feature of PDAC, their distribution is not entirely uniform. Instead, a distinct enrichment of TP53 alterations is observed in gemcitabine-treated early-onset PDAC, whereas late-onset disease exhibits a more stable mutation pattern across treatment contexts. This suggests that TP53-driven tumor biology may be differentially shaped by age and therapeutic exposure, with potential implications for treatment response and disease progression.

#### 3.3.2 PI3K gene-level landscape suggests treatment-associated diversification in late-onset PDAC

At the gene level, the PI3K pathway displayed a markedly lower overall mutation burden than the TP53 pathway, with alterations distributed across multiple low-frequency nodes rather than dominated by a single recurrent driver. In early-onset PDAC, PI3K-related alterations were rare and restricted to gemcitabine-treated tumors, where isolated mutations in PIK3CA and RPTOR were each detected in 6.7% of cases, while no alterations were observed in the non-gemcitabine-treated early-onset subgroup (Table S5). No mutations were identified in PTEN, PIK3R1, PIK3R2, PIK3R3, INPP4B, AKT1/2/3, PPP2R1A, TSC1, TSC2, STK11, RHEB, RICTOR, or MTOR in early-onset untreated tumors, underscoring the sparsity of PI3K pathway disruption in this setting.

A broader and more heterogeneous PI3K mutational spectrum was observed in late-onset PDAC, particularly among gemcitabine-treated patients. In this subgroup, PIK3CA mutations were the most frequent PI3K-axis event (4.4%), followed by lower-frequency alterations in PPP2R1A, TSC2, STK11, and RICTOR (each 2.2%), with additional sporadic events in PIK3R1, PIK3R2, PIK3R3, INPP4B, AKT2, AKT3, TSC1, MTOR, and RPTOR (Table S6). By contrast, non-treated late-onset tumors showed a substantially narrower landscape, with only STK11 mutations identified in 2.7% of cases and no detectable alterations in most other PI3K pathway genes (Table S6). Although individual gene-level comparisons did not reach statistical significance, the accumulation of multiple low-frequency events in treated late-onset tumors supports a treatment-contextual expansion of PI3K pathway complexity.

Age-based comparisons within the gemcitabine-treated cohort further emphasized this pattern. Early-onset treated tumors harbored only isolated PIK3CA and RPTOR events, whereas late-onset treated tumors exhibited a broader distribution of mutations involving upstream regulators, catalytic components, and mTOR-complex-related genes, including PIK3R1/2/3, INPP4B, AKT2/3, PPP2R1A, TSC1/2, STK11, RICTOR, and MTOR (Table S7). Despite this greater diversity in late-onset disease, no single gene differed significantly between age groups within the gemcitabine-treated stratum. Likewise, comparison of untreated early- versus late-onset PDAC revealed minimal PI3K pathway disruption overall, with only STK11 mutations detected in a small subset of late-onset untreated tumors (2.7%) and no alterations observed in early-onset untreated cases (Table S8).

Taken together, these findings indicate that PI3K pathway dysregulation in PDAC is not characterized by a single dominant gene-level event, but rather by low-frequency alterations across multiple signaling components. This pattern is most evident in gemcitabine-treated late-onset PDAC, where the pathway appears more genomically diversified than in early-onset or untreated disease, suggesting that therapeutic exposure may coincide with broader PI3K-axis molecular heterogeneity.

#### 3.3.3 Integrated interpretation

Taken together, gene-level analyses reveal that the relative consistency observed at the pathway level (Section 3.2) conceals important age- and treatment-dependent differences in the underlying molecular architecture of PDAC. The TP53 axis is characterized by a dominant, recurrent alteration pattern driven primarily by TP53 itself, with a notable enrichment in gemcitabine-treated early-onset tumors, suggesting a potential interaction between tumor suppressor dysfunction and therapeutic exposure in this subgroup. In contrast, the PI3K pathway exhibits a fundamentally different structure, marked by low-frequency alterations distributed across multiple genes rather than a single dominant driver. This dispersed pattern is most evident in gemcitabine-treated late-onset PDAC, where an expanded spectrum of PI3K-related alterations, including components of upstream regulation, catalytic signaling, and mTOR complex activity, emerges. Conversely, early-onset and untreated tumors display minimal PI3K pathway disruption, indicating a more constrained signaling landscape in these contexts.

Overall, these findings highlight two distinct modes of pathway dysregulation in PDAC: a TP53-driven architecture that is broadly conserved but modulated by age and treatment, and a PI3K pathway that appears more plastic and context-dependent, with evidence of increased molecular diversification in association with gemcitabine exposure.

### 3.4 Context-Specific Survival Impact of TP53 Pathway Alterations

To determine the prognostic implications of TP53 pathway dysregulation, we performed Kaplan-Meier survival analyses across PDAC subgroups stratified by age at diagnosis and gemcitabine exposure (Figure 1a-d).

**Figure 1.**
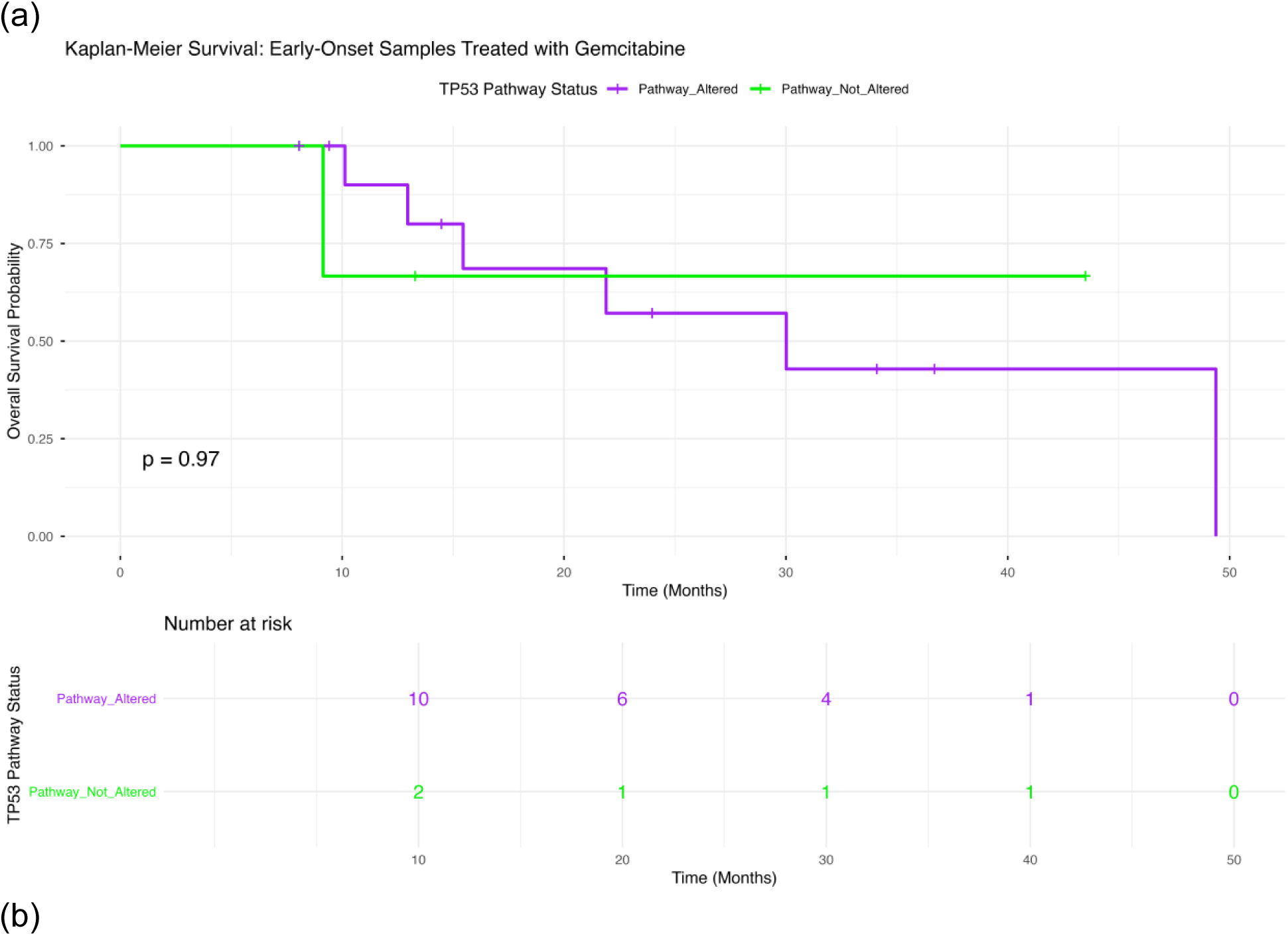
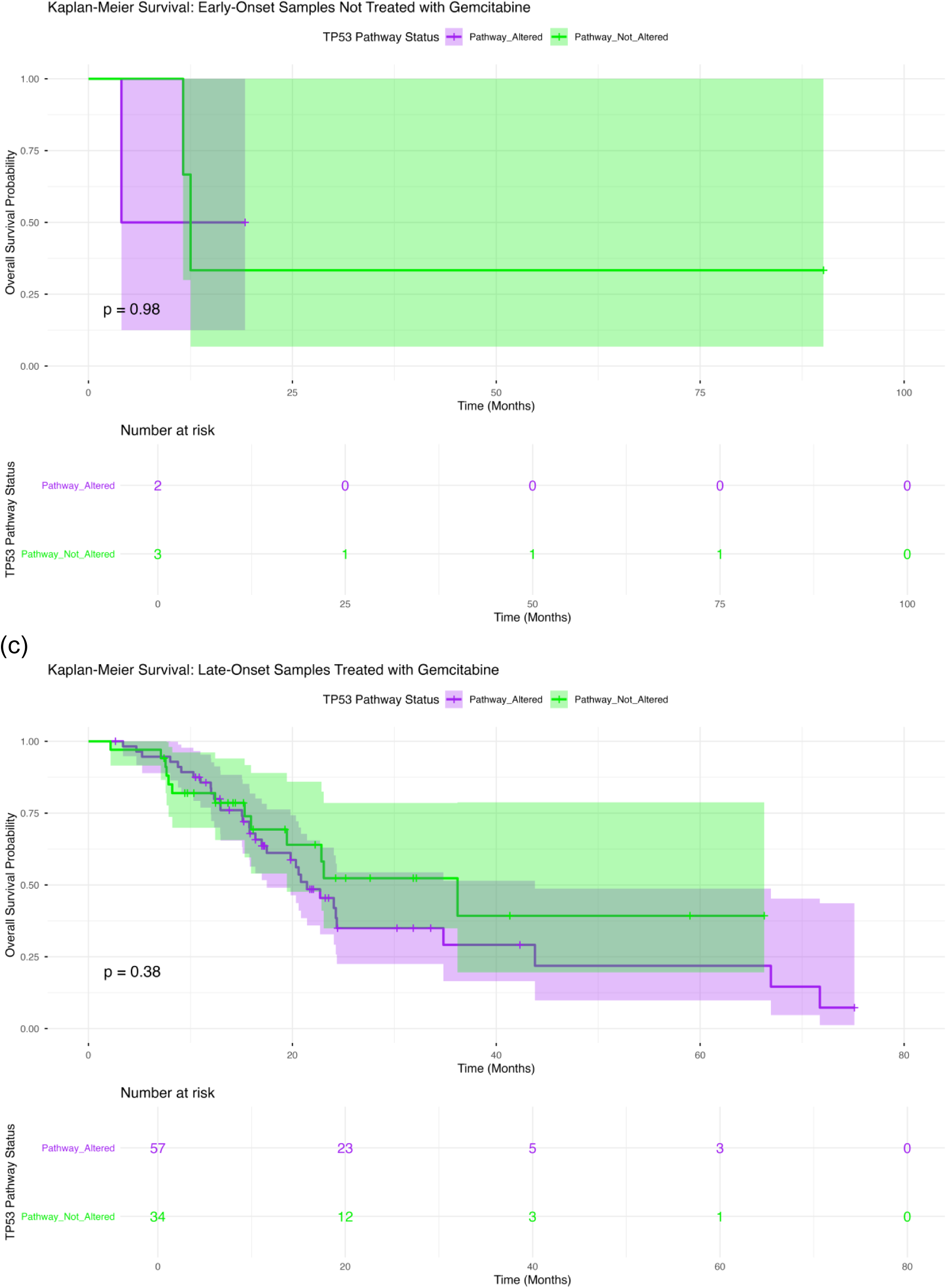
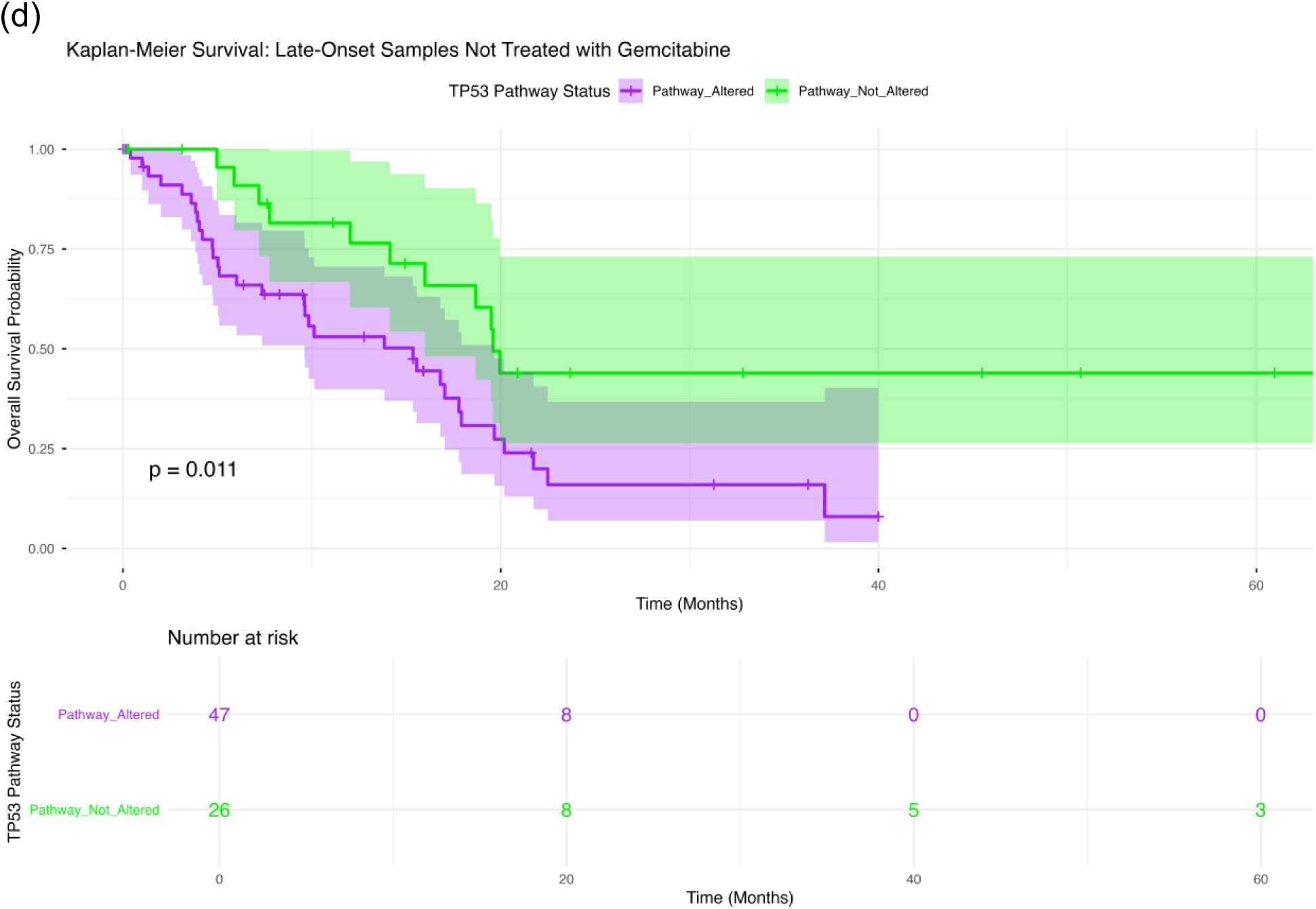
Overall survival stratified by TP53 pathway alteration status in age- and treatment-defined PDAC subgroups. Kaplan-Meier analyses illustrate differences in overall survival between patients with TP53 pathway-altered tumors and those lacking detectable TP53 alterations, with stratification based on age at diagnosis and gemcitabine exposure. The four panels represent: (a) early-onset PDAC (<50 years) receiving gemcitabine, (b) early-onset PDAC without gemcitabine treatment, (c) late-onset PDAC (≥50 years) receiving gemcitabine, and (d) late-onset PDAC without gemcitabine exposure. Confidence intervals (95%) are shown as shaded regions surrounding survival curves, and corresponding numbers at risk are provided below each panel.

#### 3.4.1 Early-Onset PDAC Under Gemcitabine Exposure

Among early-onset patients treated with gemcitabine (Figure 1a), overall survival was comparable between tumors harboring TP53 pathway alterations and those without detectable alterations (p = 0.97). Survival curves closely overlapped across the duration of follow-up, suggesting no discernible prognostic impact of TP53 status in this subgroup. The relatively small sample size contributed to broad confidence intervals, limiting sensitivity to detect subtle differences.

#### 3.4.2 Early-Onset PDAC Without Gemcitabine Exposure

In early-onset patients who did not receive gemcitabine (Figure 1b), TP53 pathway status similarly showed no association with overall survival (p = 0.98). Although slight variability in survival patterns was observed, no consistent separation between groups was evident. These findings should be interpreted cautiously given the limited number of patients and events.

#### 3.4.3 Late-Onset PDAC Under Gemcitabine Exposure

For late-onset PDAC patients undergoing gemcitabine-based therapy (Figure 1c), survival outcomes between TP53-altered and non-altered tumors demonstrated a modest degree of divergence; however, this did not achieve statistical significance (p = 0.38). While early survival was comparable, a gradual decline in survival probability was observed among TP53-altered cases at later time points. Nonetheless, overlapping confidence intervals indicate that these differences are not definitive.

#### 3.4.4 Late-Onset PDAC Without Gemcitabine Exposure

A distinct pattern was observed in late-onset patients not treated with gemcitabine (Figure 1d), where TP53 pathway status was significantly associated with survival outcomes (p = 0.011). Patients without TP53 alterations experienced improved overall survival compared to those with TP53 pathway mutations. The divergence between curves occurred early and persisted throughout follow-up, indicating a sustained adverse prognostic effect of TP53 alterations in this clinical context.

In summary, the impact of TP53 pathway alterations on survival in PDAC appears to depend on both age and treatment exposure. While no significant association was observed in early-onset disease or in gemcitabine-treated patients, TP53 alterations identify a subgroup with worse prognosis among late-onset, untreated patients. These findings underscore the importance of integrating molecular alterations with clinical stratification to refine prognostic assessment in PDAC.

### 3.5 Context-Specific Survival Impact of PI3K Pathway Alterations

We investigated the association between PI3K pathway alterations and overall survival across PDAC subgroups defined by age at diagnosis and gemcitabine exposure using Kaplan-Meier analyses (Figure 2a-d).

**Figure 2.**
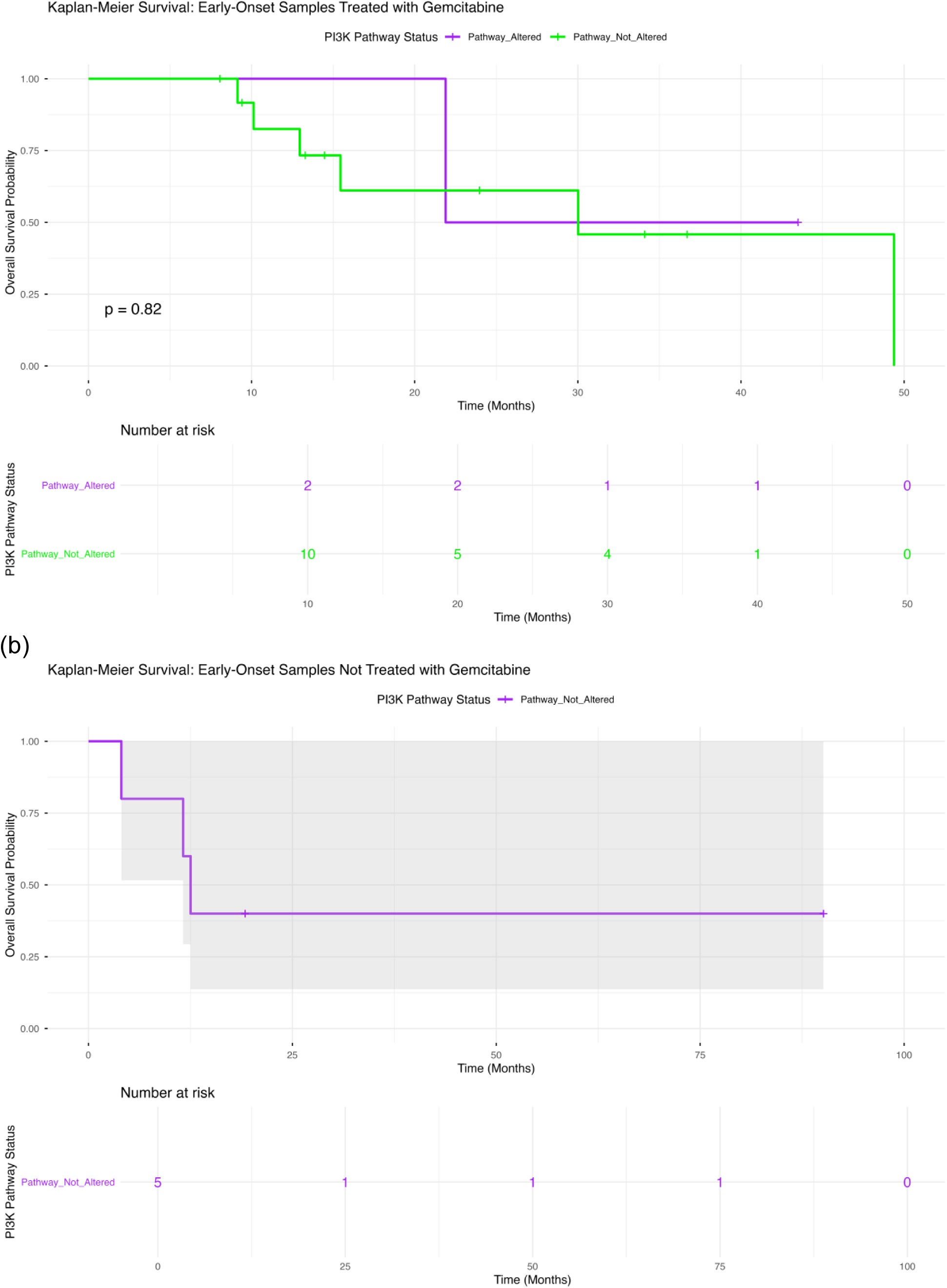
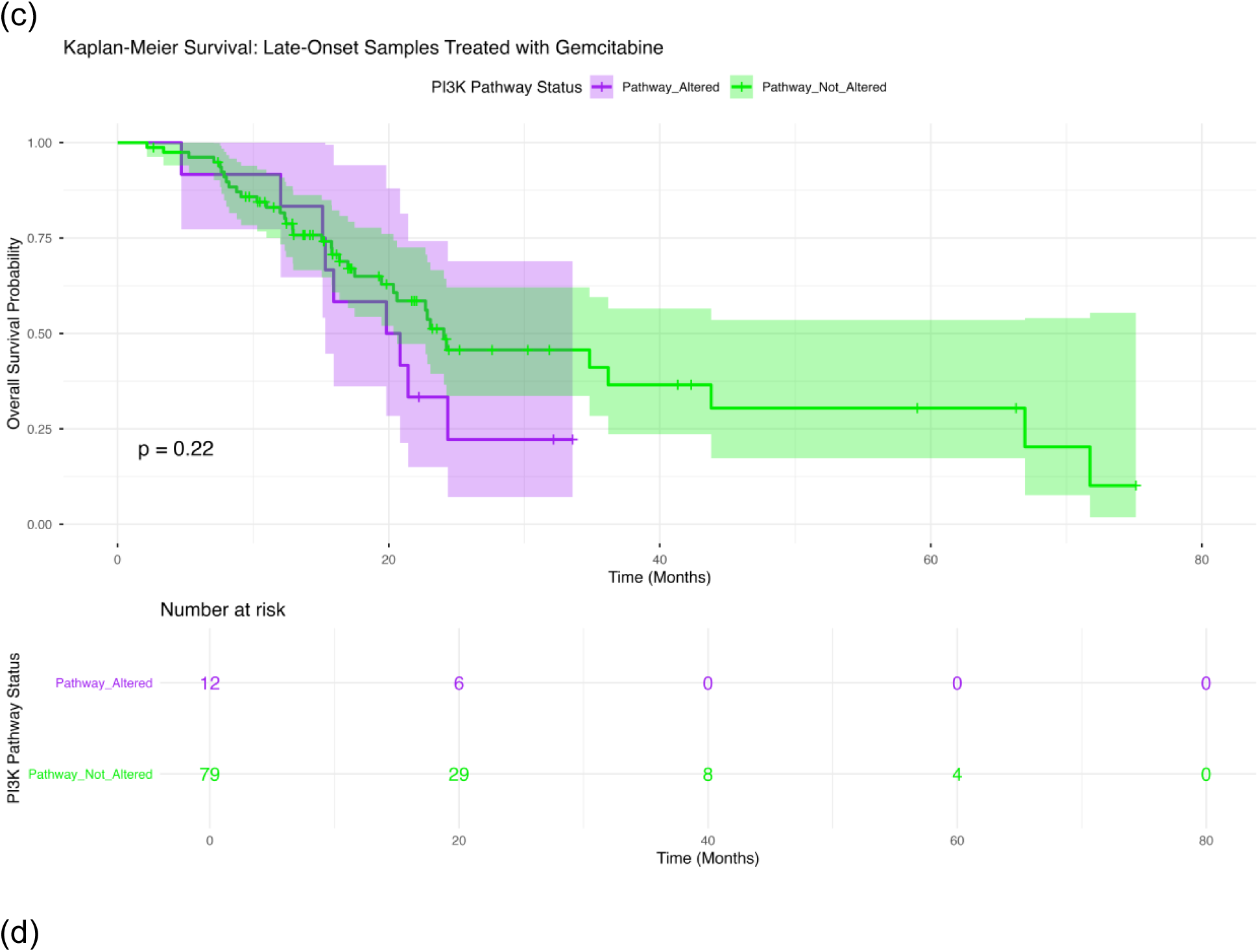
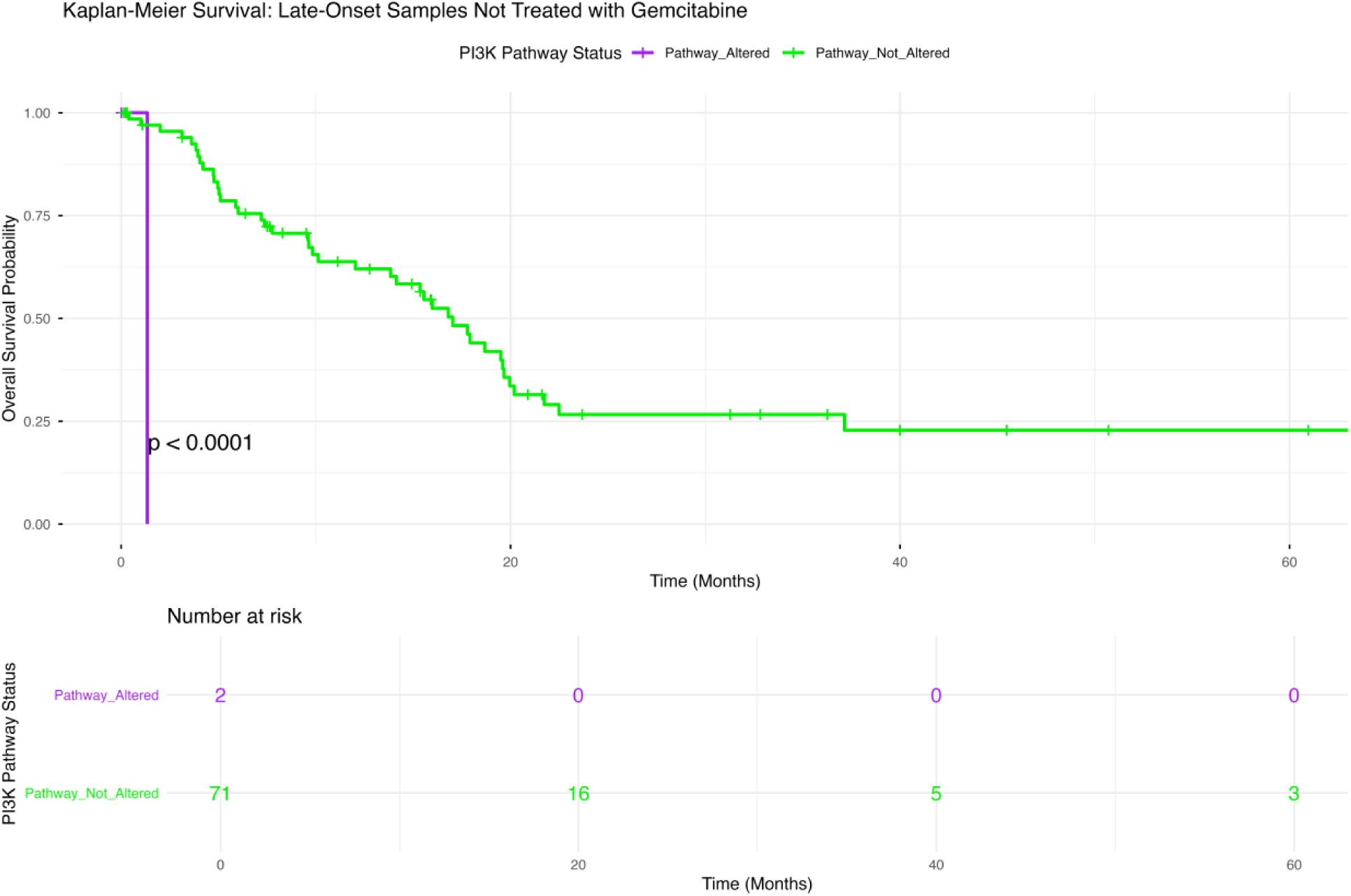
Overall survival stratified by PI3K pathway alteration status in age- and treatment-defined PDAC subgroups. Kaplan-Meier analyses compare survival outcomes between patients with PI3K pathway-altered tumors and those without detectable alterations, with stratification based on age at diagnosis and gemcitabine exposure. The panels represent: (a) early-onset PDAC (<50 years) receiving gemcitabine, (b) early-onset PDAC without gemcitabine exposure, (c) late-onset PDAC (≥50 years) receiving gemcitabine, and (d) late-onset PDAC without gemcitabine treatment. Within each subgroup, survival trajectories for PI3K-altered and PI3K-wild-type tumors are evaluated. Shaded regions denote 95% confidence intervals, and risk tables beneath each plot indicate the number of patients at risk over time, supporting interpretation of survival dynamics across follow-up.

#### 3.5.1 Early-Onset PDAC with Gemcitabine Exposure

In early-onset patients receiving gemcitabine (Figure 2a), overall survival was comparable between PI3K-altered and PI3K-wild-type tumors (p = 0.82). The survival trajectories for both groups were largely overlapping throughout follow-up, with no consistent divergence observed. Interpretation is limited by the small number of cases harboring PI3K alterations, resulting in broad confidence intervals.

#### 3.5.2 Early-Onset PDAC without Gemcitabine Exposure

For early-onset patients not treated with gemcitabine (Figure 2b), meaningful comparison by PI3K pathway status was not possible, as no PI3K alterations were detected in this subgroup. Consequently, survival estimates reflect only the PI3K-wild-type population, preventing assessment of differential outcomes.

#### 3.5.3 Late-Onset PDAC with Gemcitabine Exposure

Among late-onset patients treated with gemcitabine (Figure 2c), survival outcomes did not significantly differ according to PI3K pathway status (p = 0.22). Although PI3K-altered tumors showed a tendency toward lower survival probabilities at earlier time points, this pattern was not sustained, and the survival curves converged with substantial overlap in confidence intervals.

#### 3.5.4 Late-Onset PDAC without Gemcitabine Exposure

In contrast, late-onset patients who did not receive gemcitabine (Figure 2d) exhibited a pronounced survival difference based on PI3K pathway status (p < 0.0001). Patients with PI3K pathway alterations had markedly reduced overall survival compared to those without alterations. The separation between survival curves occurred early and persisted over time, indicating a robust and sustained adverse prognostic effect.

Analysis revealed a context-dependent role of PI3K pathway alterations in PDAC survival. No significant effect was observed in early-onset or gemcitabine-treated patients, whereas PI3K alterations delineated a high-risk subgroup among untreated late-onset patients. These findings support the integration of pathway-level information with clinical variables to refine prognostic models and guide precision oncology.

### 3.6 AI-Enabled Identification of Context-Specific Molecular Patterns

#### 3.6.1 AI-HOPE-TP53: Context-Aware Cohort Modeling and Pathway Interrogation

To extend conventional pathway analyses, we utilized the AI-HOPE-TP53 framework to perform structured, natural language-driven exploratory analyses within the integrated PDAC clinical-genomic dataset. This approach enabled flexible cohort construction, real-time survival evaluation, and targeted enrichment testing under explicitly defined clinical and molecular conditions.

As an initial demonstration (Figure S1), the AI agent was tasked with constructing clinically matched cohorts based on age and treatment exposure and evaluating survival outcomes according to TP53 pathway status. Using user-defined criteria, the system identified late-onset PDAC patients not treated with gemcitabine and partitioned them into TP53 pathway-altered (n = 47) and non-altered (n = 26) groups. Kaplan-Meier analysis revealed a statistically significant difference in overall survival between these cohorts (log-rank p = 0.0113), with TP53 pathway alterations associated with inferior outcomes. This example highlights the ability of conversational AI to reproducibly generate clinically relevant subgroups and perform survival modeling within narrowly defined contexts.

We next applied the AI-HOPE-TP53 agent to investigate the relationship between gene-level TP53 mutations and broader pathway-level dysregulation (Figure S2). Restricting the analysis to tumors harboring TP53 mutations, the system constructed comparison cohorts based on the presence or absence of additional TP53 pathway alterations. Contingency-based statistical testing demonstrated a highly significant enrichment of pathway-level alterations among TP53-mutant tumors (p ≈ 0), with a strong odds ratio indicating substantial overlap between mutation status and pathway activation. This analysis underscores the dominant role of TP53 as the central node within its pathway and illustrates how conversational AI can quantify hierarchical relationships between gene-level events and pathway-level architecture.

AI-driven exploratory analyses highlight the utility of conversational frameworks for interrogating clinically stratified datasets. Through dynamic cohort definition, integrated survival analysis, and enrichment testing, AI-HOPE-TP53 enables efficient hypothesis generation and validation across complex clinical and molecular contexts.

#### 3.6.2 AI-HOPE-PI3K: Contextual Analysis of PI3K Pathway Associations

We next applied the AI-HOPE-PI3K agent to systematically explore clinical and genomic correlates of PI3K pathway alterations across the PDAC cohort, with particular emphasis on treatment-associated patterns and pathway-level interactions (Figures S3-S4).

To establish a global view of PI3K pathway-associated features, the agent first constructed cohorts stratified by PI3K pathway status (Figure S3). Tumors harboring PI3K pathway alterations (n = 16) were compared with a substantially larger group lacking such alterations (n = 168), reflecting the relatively low prevalence of PI3K dysregulation in this dataset. Through automated Chi-square-based association testing, the AI framework identified significant relationships between PI3K pathway status and multiple components of the PI3K signaling axis, including PIK3CA, STK11, MTOR, PPP2R1A, RICTOR, TSC2, and RPTOR. These findings indicate coordinated perturbation across upstream regulators and mTOR complex-related nodes rather than reliance on a single dominant alteration.

Beyond canonical pathway members, the analysis also revealed associations with additional signaling and regulatory genes, including NLK, RNF43, and TCF7, as well as calcium channel-related genes within the CACNA family. Enrichment of WNT pathway alterations further suggests cross-talk between PI3K signaling and other oncogenic networks. Clinically, PI3K pathway status was associated with gemcitabine treatment exposure and progression-free status, and differences were also observed across histologic subtypes. Together, these results define PI3K-altered PDAC as a distinct molecular subset characterized by multi-node pathway involvement and broader signaling network integration.

We then investigated whether PI3K pathway alterations were preferentially associated with gemcitabine exposure using an AI-driven contingency analysis (Figure S4). The agent compared treatment distribution between PI3K-altered and PI3K-wild-type tumors, demonstrating a significantly higher proportion of gemcitabine-treated cases among PI3K-altered tumors (87.5% vs. 54.76%). Statistical testing confirmed this association (p = 0.023), with an elevated odds ratio indicating enrichment of PI3K pathway alterations in the treatment-exposed population.

This finding supports a model in which PI3K pathway dysregulation may be linked to treatment context, potentially reflecting therapy-associated selection or adaptive signaling responses. Importantly, the AI-HOPE-PI3K framework enabled rapid identification of this relationship through structured query execution and automated statistical testing, highlighting its utility for uncovering clinically relevant, treatment-associated molecular patterns.

PI3K pathway alterations in PDAC exhibit both genomic heterogeneity and context-dependent associations with treatment exposure and signaling networks. The AI-driven approach enables efficient integration of molecular and clinical data, offering insights into treatment-dependent disease biology.

## 4. Discussion

PDAC remains a biologically aggressive and clinically heterogeneous malignancy in which treatment response is shaped by both intrinsic tumor genetics and the context in which those alterations operate. In this study, we applied a conversational artificial intelligence strategy, using AI-HOPE-TP53 (33) and AI-HOPE-PI3K (34), to interrogate TP53- and PI3K-centered pathway architecture across a clinically stratified PDAC cohort defined by age at diagnosis and gemcitabine exposure. Several key findings emerged from this analysis. First, TP53 pathway disruption was common across the cohort, but gene-level analysis revealed that TP53 mutations were particularly enriched in gemcitabine-treated early-onset PDAC. Second, PI3K pathway alterations were much less frequent overall, yet they were significantly enriched in gemcitabine-treated late-onset PDAC and displayed a broader, more distributed mutational architecture in that setting. Third, the prognostic value of both pathways was highly context dependent, with the clearest survival differences observed among late-onset patients who had not received gemcitabine, where the absence of either TP53 or PI3K pathway alterations identified more favorable-outcome subgroups.

A central message of this work is that pathway prevalence alone does not fully capture clinically meaningful biology. At the aggregate level, TP53 pathway alterations appeared broadly stable across age and treatment categories, which could suggest that TP53 dysfunction is simply a ubiquitous background feature of PDAC. However, deeper inspection at the gene level showed that this apparent stability conceals meaningful differences in the distribution of the dominant event within the pathway, namely TP53 mutation itself. In particular, the significantly higher TP53 mutation frequency in gemcitabine-treated early-onset tumors relative to gemcitabine-treated late-onset tumors suggests that early-onset PDAC may rely more heavily on TP53-driven biology under chemotherapy-associated selective pressure. This does not necessarily imply that gemcitabine induces TP53 alterations, but it raises the possibility that tumors entering or persisting through gemcitabine exposure in younger patients may be enriched for TP53-disrupted disease states that confer enhanced fitness, genomic instability, or resistance-associated phenotypes.

This observation is biologically plausible. TP53 loss is tightly linked to impaired apoptosis, defective cell-cycle checkpoint control, and increased tolerance of genotoxic stress, all of which may influence response to cytotoxic therapy. In PDAC, where treatment resistance is often multifactorial, higher TP53 mutation rates in gemcitabine-treated early-onset tumors may reflect a disease subset better equipped to survive DNA damage and metabolic stress. The trend toward higher TP53 mutation frequency in treated versus untreated early-onset disease, although not statistically significant, further supports this interpretation and suggests that a larger cohort may reveal a more robust treatment-associated signal in younger patients. By contrast, late-onset disease showed similar TP53 mutation frequencies regardless of gemcitabine exposure, implying that TP53-driven biology in older patients may be more constitutive and less specifically shaped by this treatment context.

The PI3K pathway showed a very different pattern. Unlike TP53, PI3K pathway alterations were uncommon at the cohort level, but their distribution was more clinically informative. The significant enrichment of PI3K pathway alterations in gemcitabine-treated late-onset PDAC points to a treatment-contextual expansion of this signaling axis in older patients. Importantly, this enrichment was not driven by a single dominant gene. Rather, the pathway in treated late-onset tumors was characterized by low-frequency alterations spread across multiple nodes, including PIK3CA, PIK3R family members, AKT isoforms, TSC1/2, STK11, RICTOR, MTOR, and related regulators. This kind of dispersed architecture suggests that PI3K pathway activation in PDAC may arise through several parallel or partially redundant molecular routes, especially under therapeutic pressure.

That pattern is conceptually important. A pathway dominated by one highly recurrent event often implies a central, shared dependency. In contrast, a pathway altered through many different low-frequency lesions may reflect adaptive plasticity rather than a single essential driver. In this study, the broader mutational distribution seen in gemcitabine-treated late-onset PDAC is consistent with the idea that PI3K signaling may serve as a flexible escape route in the setting of treatment exposure. Even though individual PI3K-axis genes did not show statistically significant differences on their own, the collective diversification of the pathway in treated late-onset disease is difficult to dismiss as random noise, particularly because the pathway-level enrichment was significant. This may reflect subclonal selection, compensatory survival signaling, or metabolic rewiring associated with chemotherapy tolerance.

Another important finding is that the prognostic relevance of both pathways was concentrated in late-onset patients who did not receive gemcitabine. In that subgroup, absence of TP53 pathway alterations was associated with significantly better overall survival, and the same was true, even more strongly, for the absence of PI3K pathway alterations. By contrast, in gemcitabine-treated patients and in early-onset disease, neither pathway consistently stratified survival. This suggests that the prognostic meaning of pathway status is not fixed, but instead depends on treatment exposure and disease context.

Several explanations may account for this pattern. In untreated late-onset disease, pathway alterations may more directly reflect the baseline natural history of the tumor, unmodified by the confounding effects of systemic therapy. In that setting, tumors lacking TP53 or PI3K pathway disruption may represent biologically less aggressive subsets or may rely on alternate oncogenic programs associated with slower progression. Conversely, once patients are exposed to gemcitabine, treatment-related selection, line-of-therapy effects, and unmeasured clinical confounders may dilute or obscure the prognostic signal attributable to any single pathway. This may be especially relevant in PDAC, where treatment decisions often correlate with fitness, stage, comorbidity, and other factors that are incompletely captured in retrospective clinicogenomic datasets.

The survival findings also reinforce a broader conceptual point: prognostic markers and predictive markers are not interchangeable. The enrichment of PI3K pathway alterations in gemcitabine-treated late-onset PDAC does not automatically mean PI3K status predicts gemcitabine response, just as the adverse survival associated with TP53 or PI3K alterations in untreated late-onset disease does not necessarily establish those pathways as treatment-selection biomarkers. Rather, the present data indicate that these pathways carry different forms of clinical information depending on setting. In some contexts they appear to track tumor aggressiveness, while in others they may reflect treatment-associated evolutionary pressure. Distinguishing those roles will require datasets with more granular therapy timing, response data, and multivariable modeling.

Methodologically, this study also demonstrates the practical value of conversational artificial intelligence as an analytic interface for precision oncology. AI-HOPE-TP53 and AI-HOPE-PI3K enabled rapid cohort construction, stratified pathway interrogation, and hypothesis generation across multiple overlapping clinical dimensions without abandoning reproducibility. This is particularly useful in studies like this one, where clinically relevant subgrouping is not simple. Investigators may need to move repeatedly between age-defined, treatment-defined, pathway-level, gene-level, and survival-based analyses. Conversational AI does not replace statistical rigor, but it can substantially reduce friction in exploratory and semi-structured analysis, allowing complex questions to be posed and iteratively refined in ways that are more aligned with translational reasoning. The fact that AI-derived findings were validated using conventional statistical approaches strengthens the case that such systems can serve as a credible analytic layer rather than a black-box shortcut.

These results should be interpreted in light of several limitations. The most important is subgroup size, especially in early-onset PDAC and particularly in early-onset patients not treated with gemcitabine. Small numbers reduce power, widen confidence intervals, and make both gene-level and survival analyses vulnerable to instability. Second, gemcitabine exposure was handled as a binary variable, whereas real-world treatment is more nuanced and includes combination regimens, treatment sequencing, dose intensity, treatment duration, and variable temporal relationships to sequencing. Third, the cohort was dominated by stage II disease, which may limit generalizability to more advanced PDAC populations. Fourth, DNA-level pathway alteration status does not directly measure pathway activity. A tumor without a mutation in a PI3K pathway gene may still have functional pathway activation through epigenetic, transcriptional, post-translational, stromal, or metabolic mechanisms. Similarly, not all mutations within a pathway are functionally equivalent. Fifth, the demographic composition of the cohort limits equity-oriented interpretation. Hispanic/Latino cases were sparse and ethnicity was unknown in a sizable fraction of patients, preventing meaningful evaluation of ancestry-or ethnicity-associated differences in TP53 or PI3K biology.

Future work should build on these observations in several directions. Validation in larger and independent PDAC cohorts is essential, particularly to confirm the enrichment of TP53 mutations in gemcitabine-treated early-onset disease and the treatment-associated diversification of PI3K signaling in late-onset PDAC. More detailed treatment annotation will help determine whether these are truly chemotherapy-associated features or reflections of broader disease selection. Functional studies are also needed. For TP53, this means testing whether early-onset, TP53-mutant PDAC shows distinct apoptotic thresholds, genomic instability profiles, or chemotherapy adaptation states. For PI3K, it will be important to determine whether the distributed set of low-frequency alterations identified in treated late-onset tumors converges on shared downstream signaling outputs that could still be therapeutically targetable. Integration of transcriptomic, phosphoproteomic, spatial, and microenvironmental data would be especially informative, because these layers may reveal whether DNA alterations translate into pathway activation states that matter clinically.

In summary, this study supports a model in which TP53 and PI3K pathways contribute to PDAC biology in distinct but clinically meaningful ways. TP53 alterations form a dominant and recurrent backbone of disease, but their enrichment in gemcitabine-treated early-onset PDAC suggests that age and treatment can shape even highly recurrent tumor suppressor events. PI3K alterations, in contrast, are infrequent but more plastic, and their significant enrichment in gemcitabine-treated late-onset tumors points to a context in which signaling diversity may expand under treatment pressure. The strongest prognostic effects for both pathways were seen in late-onset patients not treated with gemcitabine, indicating that pathway status can stratify survival most clearly when not obscured by treatment-related confounding. Together, these findings argue for moving beyond simple pathway-present versus pathway-absent classification and toward clinically contextualized, gene-level dependency mapping. They also illustrate how conversational AI can accelerate that process by making complex, multidimensional molecular analyses more accessible, reproducible, and hypothesis-generating within a precision oncology framework.

## 5. Conclusions

In this clinically stratified PDAC cohort, TP53 and PI3K pathways showed distinct patterns of molecular organization and clinical relevance. TP53 pathway disruption was common across the cohort, but TP53 mutations were particularly enriched in gemcitabine-treated early-onset PDAC. In contrast, PI3K pathway alterations were significantly enriched in gemcitabine-treated late-onset disease and were characterized by a broader, low-frequency mutational spectrum distributed across multiple pathway components. Survival analyses further showed that the absence of TP53 or PI3K pathway alterations identified favorable-prognosis subsets within late-onset PDAC patients who did not receive gemcitabine.

These findings indicate that age at onset and treatment exposure shape the biological meaning of pathway alterations in PDAC. They also highlight the value of conversational artificial intelligence as a practical platform for context-aware, pathway-centric interrogation of clinicogenomic data. By enabling rapid and reproducible analysis of clinically stratified molecular subgroups, AI-HOPE-TP53 and AI-HOPE-PI3K support a more nuanced precision oncology approach to this highly lethal disease.

## Data Availability

All data used in the present study is publicly available at https://www.cbioportal.org/. The datasets used in our study were aggregated/summary data, and no individual-level data were used. Additional data can be provided upon reasonable request to the authors.

## Supplementary Materials

**Table S1.** Comparison of Early-Onset PDAC Patients Treated with Gemcitabine Versus Those Not Treated with Gemcitabine.

| TP53 Pathway |  |  |  |
| --- | --- | --- | --- |
| Gene | Early-Onset<br>Treated with Gemcitabine<br>n (%) | Early-Onset<br>Not Treated with<br>Gemcitabine<br>n (%) | p-value |
| TP53 Mutation |  |  |  |
| Present | 13 (86.7%) | 2 (40.0%) | 0.07263 |
| Absent | 2 (13.3%) | 3 (60.0%) |  |
| MDM2 Mutation |  |  |  |
| Present | 0 (0.0%) | 0 (0.0%) | 1 |
| Absent | 15 (100.0%) | 5 (100.0%) |  |
| MDM4 Mutation |  |  |  |
| Present | 0 (0.0%) | 0 (0.0%) | 1 |
| Absent | 15 (100.0%) | 5 (100.0%) |  |
| CDKN1A Mutation |  |  |  |
| Present | 0 (0.0%) | 0 (0.0%) | 1 |
| Absent | 15 (100.0%) | 5 (100.0%) |  |
| CDKN2A Mutation |  |  |  |
| Present | 2 (13.3%) | 1 (20.0%) | 1 |
| Absent | 13 (86.7%) | 4 (80.0%) |  |
| ATM Mutation |  |  |  |
| Present | 0 (0.0%) | 0 (0.0%) | 1 |
| Absent | 15 (100.0%) | 5 (100.0%) |  |
| CHEK1 Mutation |  |  |  |
| Present | 0 (0.0%) | 0 (0.0%) | 1 |
| Absent | 15 (100.0%) | 5 (100.0%) |  |
| CHEK2 Mutation |  |  |  |
| Present | 0 (0.0%) | 0 (0.0%) | 1 |
| Absent | 15 (100.0%) | 5 (100.0%) |  |
| PTEN Mutation |  |  |  |
| Present | 0 (0.0%) | 0 (0.0%) | 1 |
| Absent | 15 (100.0%) | 5 (100.0%) |  |

| ATR Mutation |  |  |  |
| --- | --- | --- | --- |
| Present | 1 (6.7%) | 0 (0.0%) | 1 |
| Absent | 14 (93.3%) | 5 (100.0%) |  |
| BBC3 Mutation |  |  |  |
| Present | 0 (0.0%) | 0 (0.0%) | 1 |
| Absent | 15 (100.0%) | 5 (100.0%) |  |

**Table S2.**
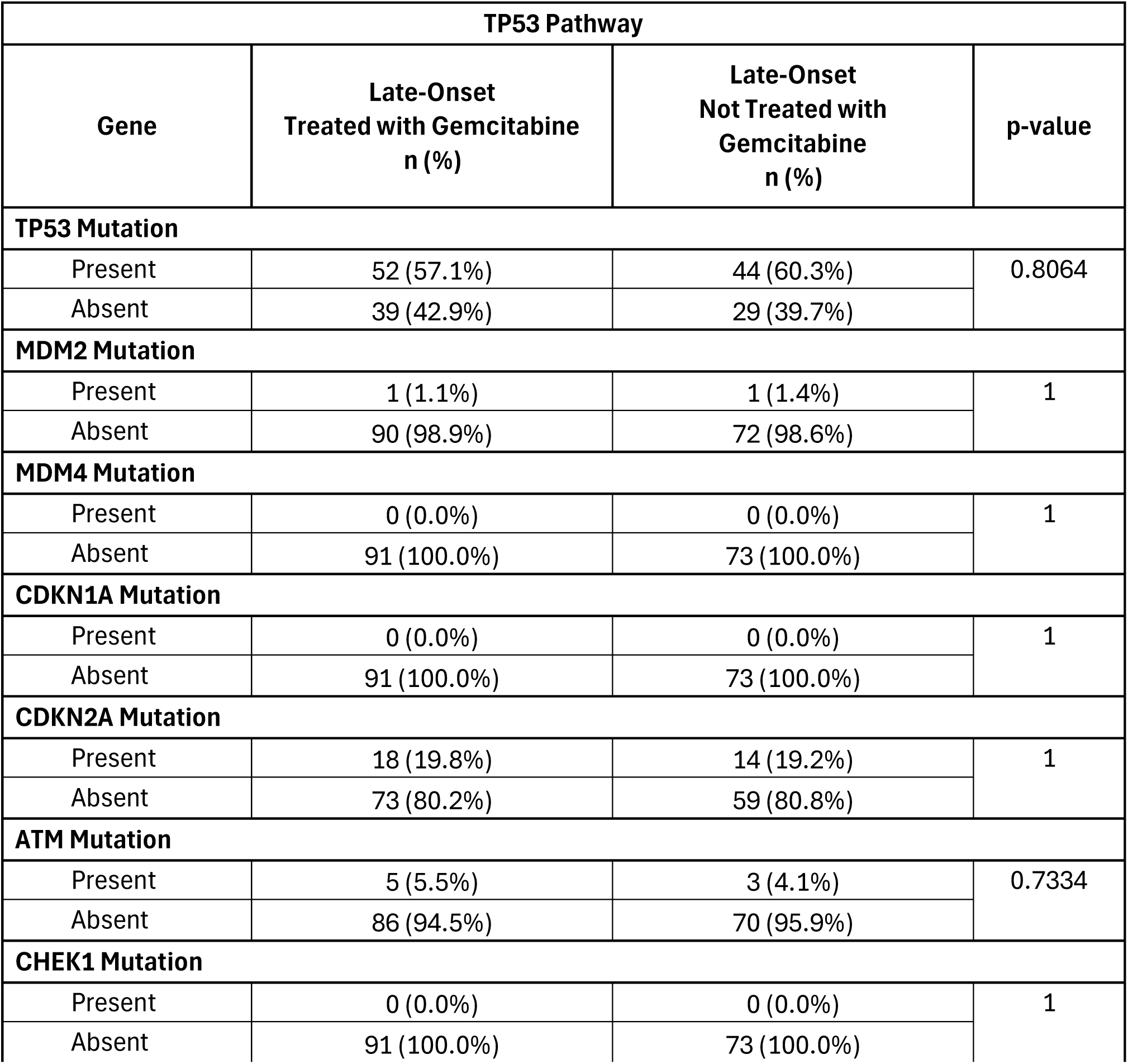
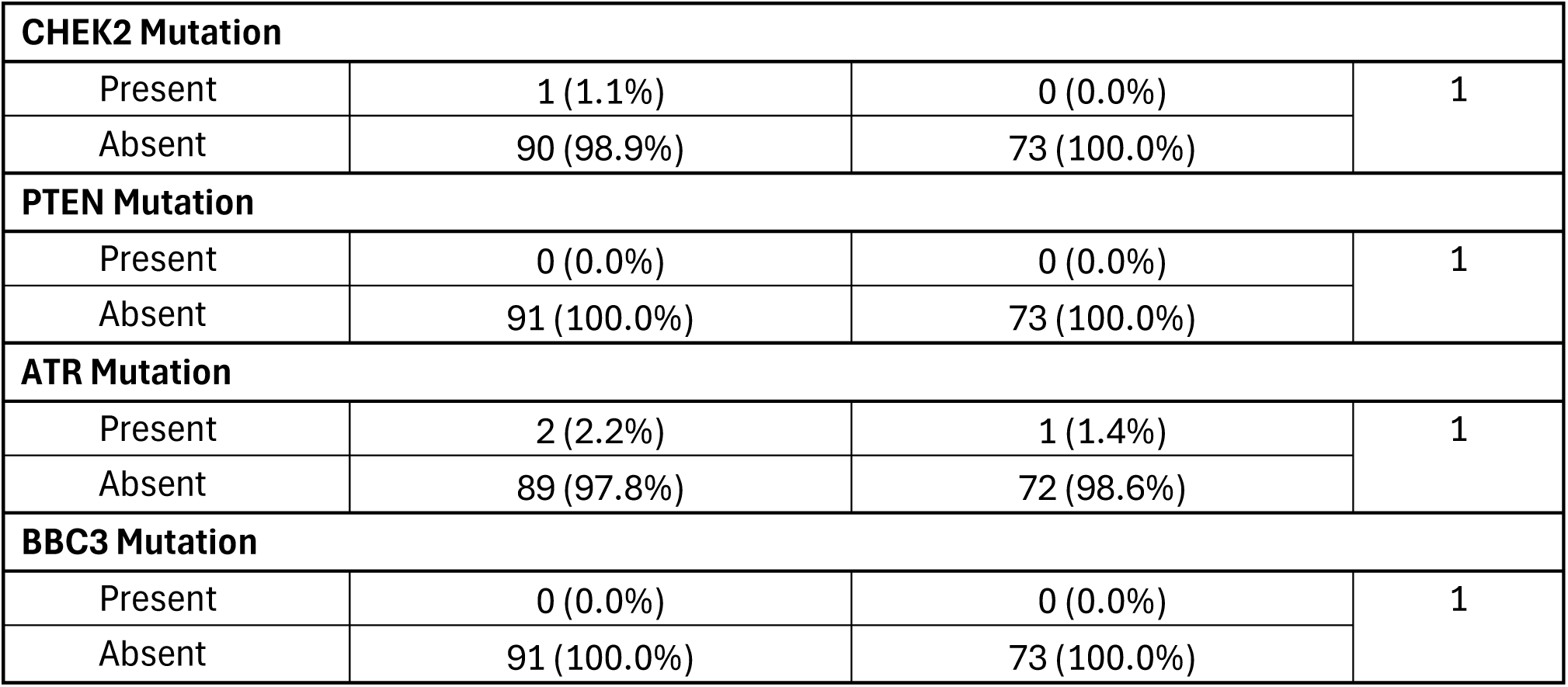
Comparison of Late-Onset PDAC Patients Treated with Gemcitabine Versus Those Not Treated with Gemcitabine.

| TP53 Pathway |  |  |  |
| --- | --- | --- | --- |
| Gene | Late-Onset<br>Treated with Gemcitabine<br>n (%) | Late-Onset<br>Not Treated with<br>Gemcitabine<br>n (%) | p-value |
| TP53 Mutation |  |  |  |
| Present | 52 (57.1%) | 44 (60.3%) | 0.8064 |
| Absent | 39 (42.9%) | 29 (39.7%) |  |
| MDM2 Mutation |  |  |  |
| Present | 1 (1.1%) | 1 (1.4%) | 1 |
| Absent | 90 (98.9%) | 72 (98.6%) |  |
| MDM4 Mutation |  |  |  |
| Present | 0 (0.0%) | 0 (0.0%) | 1 |
| Absent | 91 (100.0%) | 73 (100.0%) |  |
| CDKN1A Mutation |  |  |  |
| Present | 0 (0.0%) | 0 (0.0%) | 1 |
| Absent | 91 (100.0%) | 73 (100.0%) |  |
| CDKN2A Mutation |  |  |  |
| Present | 18 (19.8%) | 14 (19.2%) | 1 |
| Absent | 73 (80.2%) | 59 (80.8%) |  |
| ATM Mutation |  |  |  |
| Present | 5 (5.5%) | 3 (4.1%) | 0.7334 |
| Absent | 86 (94.5%) | 70 (95.9%) |  |
| CHEK1 Mutation |  |  |  |
| Present | 0 (0.0%) | 0 (0.0%) | 1 |
| Absent | 91 (100.0%) | 73 (100.0%) |  |

| CHEK2 Mutation |  |  |  |
| --- | --- | --- | --- |
| Present | 1 (1.1%) | 0 (0.0%) | 1 |
| Absent | 90 (98.9%) | 73 (100.0%) |  |
| PTEN Mutation |  |  |  |
| Present | 0 (0.0%) | 0 (0.0%) | 1 |
| Absent | 91 (100.0%) | 73 (100.0%) |  |
| ATR Mutation |  |  |  |
| Present | 2 (2.2%) | 1 (1.4%) | 1 |
| Absent | 89 (97.8%) | 72 (98.6%) |  |
| BBC3 Mutation |  |  |  |
| Present | 0 (0.0%) | 0 (0.0%) | 1 |
| Absent | 91 (100.0%) | 73 (100.0%) |  |

**Table S3.** Comparison of Early-Onset PDAC Patients Versus Late-Onset PDAC Patients Treated with Gemcitabine.

| TP53 Pathway |  |  |  |
| --- | --- | --- | --- |
| Gene | Early-Onset<br>Treated with Gemcitabine<br>n (%) | Late-Onset<br>Treated with Gemcitabine<br>n (%) | p-value |
| TP53 Mutation |  |  |  |
| Present | 13 (86.7%) | 52 (57.1%) | 0.04312 |
| Absent | 2 (13.3%) | 39 (42.9%) |  |
| MDM2 Mutation |  |  |  |
| Present | 0 (0.0%) | 1 (1.1%) | 1 |
| Absent | 15 (100.0%) | 90 (98.9%) |  |
| MDM4 Mutation |  |  |  |
| Present | 0 (0.0%) | 0 (0.0%) | 1 |
| Absent | 15 (100.0%) | 91 (100.0%) |  |
| CDKN1A Mutation |  |  |  |
| Present | 0 (0.0%) | 0 (0.0%) | 1 |
| Absent | 15 (100.0%) | 91 (100.0%) |  |
| CDKN2A Mutation |  |  |  |
| Present | 2 (13.3%) | 18 (19.8%) | 0.7312 |
| Absent | 13 (86.7%) | 73 (80.2%) |  |
| ATM Mutation |  |  |  |
| Present | 0 (0.0%) | 5 (5.5%) | 1 |
| Absent | 15 (100.0%) | 86 (94.5%) |  |
| CHEK1 Mutation |  |  |  |
| Present | 0 (0.0%) | 0 (0.0%) | 1 |
| Absent | 15 (100.0%) | 91 (100.0%) |  |
| CHEK2 Mutation |  |  |  |
| Present | 0 (0.0%) | 1 (1.1%) | 1 |
| Absent | 15 (100.0%) | 90 (98.9%) |  |
| PTEN Mutation |  |  |  |
| Present | 0 (0.0%) | 0 (0.0%) | 1 |
| Absent | 15 (100.0%) | 91 (100.0%) |  |
| ATR Mutation |  |  |  |
| Present | 1 (6.7%) | 2 (2.2%) | 0.3703 |
| Absent | 14 (93.3%) | 89 (97.8%) |  |
| BBC3 Mutation |  |  |  |
| Present | 0 (0.0%) | 0 (0.0%) | 1 |
| Absent | 15 (100.0%) | 91 (100.0%) |  |

**Table S4.**
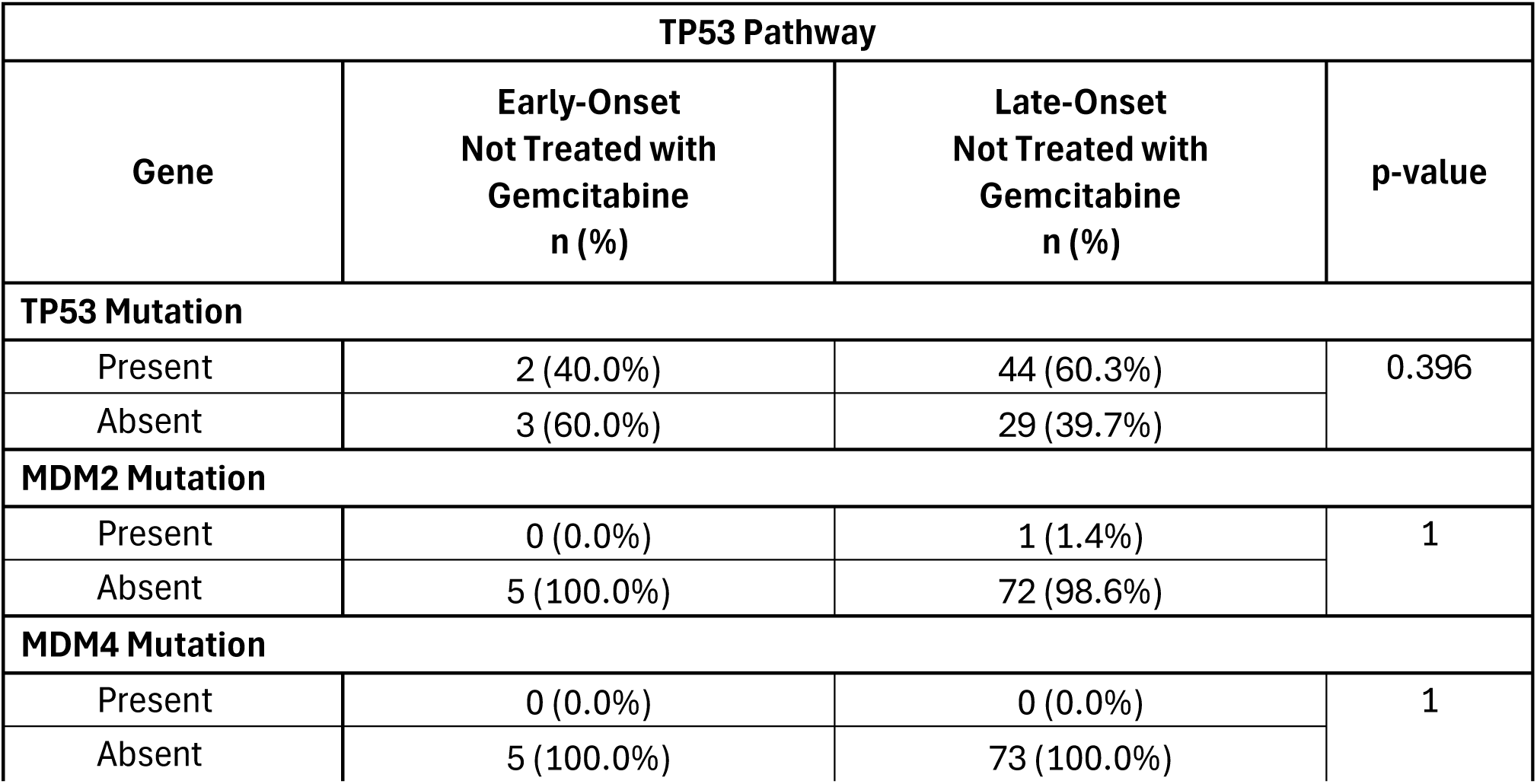
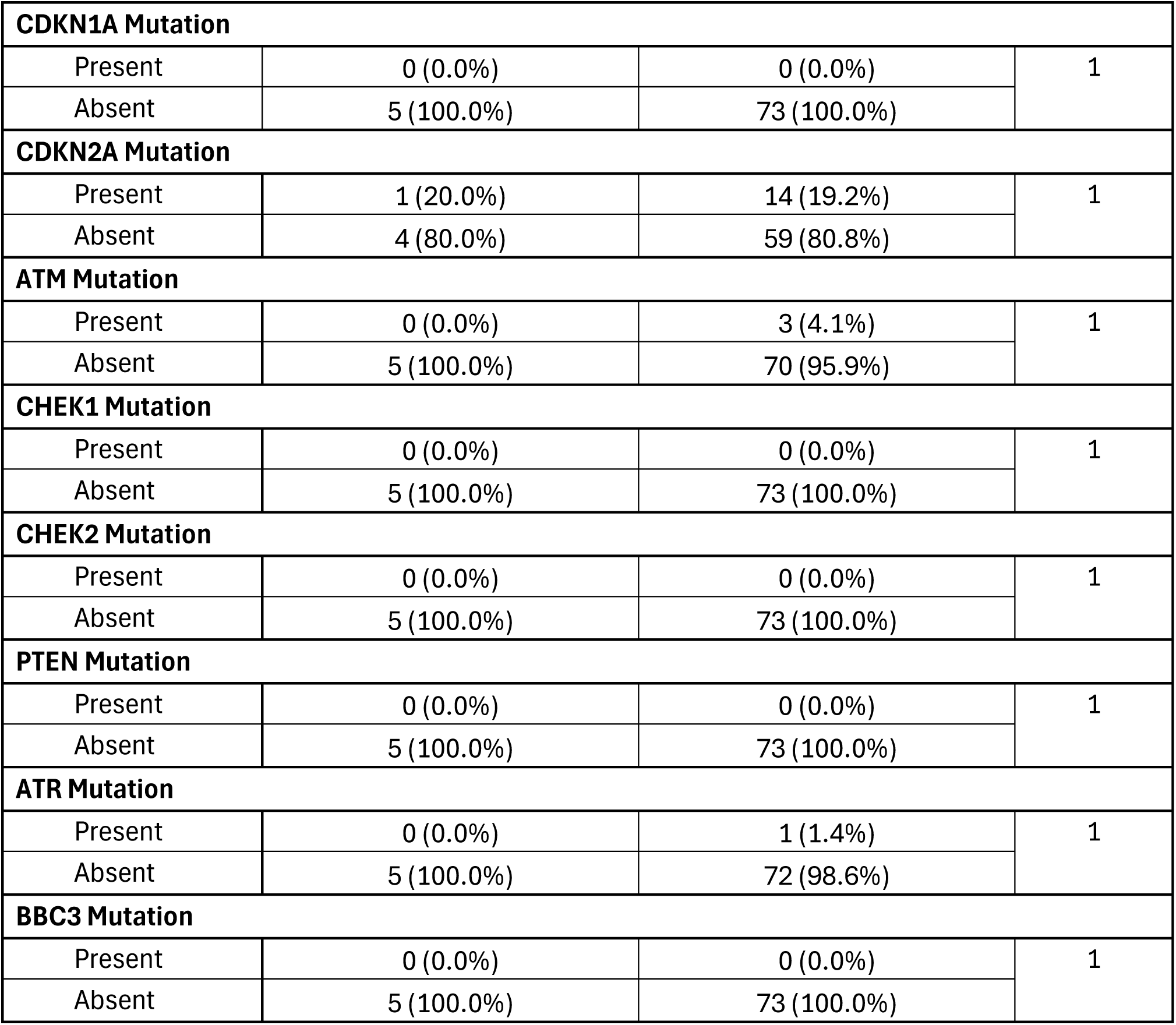
Comparison of Early-Onset PDAC Patients Versus Late-Onset PDAC Patients Not Treated with Gemcitabine.

**Table S5.** Comparison of Early-Onset PDAC Patients Treated with Gemcitabine Versus Those Not Treated with Gemcitabine.

| **PI3K Pathway** |  |  |  |
| --- | --- | --- | --- |
| **Gene** | **Early-Onset**<br>**Treated with Gemcitabine**<br>**n (%)** | **Early-Onset**<br>**Not Treated with Gemcitabine**<br>**n (%)** | **p-value** |
| **PTEN Mutation** |  |  |  |
| Present | 0 (0.0%) | 0 (0.0%) | 1 |
| Absent | 15 (100.0%) | 5 (100.0%) |  |
| PIK3R1 Mutation |  |  |  |
| Present | 0 (0.0%) | 0 (0.0%) | 1 |
| Absent | 15 (100.0%) | 5 (100.0%) |  |
| PIK3R2 Mutation |  |  |  |
| Present | 0 (0.0%) | 0 (0.0%) | 1 |
| Absent | 15 (100.0%) | 5 (100.0%) |  |
| PIK3R3 Mutation |  |  |  |
| Present | 0 (0.0%) | 0 (0.0%) | 1 |
| Absent | 15 (100.0%) | 5 (100.0%) |  |
| PIK3CA Mutation |  |  |  |
| Present | 1 (6.7%) | 0 (0.0%) | 1 |
| Absent | 14 (93.3%) | 5 (100.0%) |  |
| INPP4B Mutation |  |  |  |
| Present | 0 (0.0%) | 0 (0.0%) | 1 |
| Absent | 15 (100.0%) | 5 (100.0%) |  |
| AKT1 Mutation |  |  |  |
| Present | 0 (0.0%) | 0 (0.0%) | 1 |
| Absent | 15 (100.0%) | 5 (100.0%) |  |
| AKT2 Mutation |  |  |  |
| Present | 0 (0.0%) | 0 (0.0%) | 1 |
| Absent | 15 (100.0%) | 5 (100.0%) |  |
| AKT3 Mutation |  |  |  |
| Present | 0 (0.0%) | 0 (0.0%) | 1 |
| Absent | 15 (100.0%) | 5 (100.0%) |  |
| PPP2R1A Mutation |  |  |  |
| Present | 0 (0.0%) | 0 (0.0%) | 1 |
| Absent | 15 (100.0%) | 5 (100.0%) |  |
| TSC1 Mutation |  |  |  |
| Present | 0 (0.0%) | 0 (0.0%) | 1 |
| Absent | 15 (100.0%) | 5 (100.0%) |  |
| TSC2 Mutation |  |  |  |
| Present | 0 (0.0%) | 0 (0.0%) | 1 |
| Absent | 15 (100.0%) | 5 (100.0%) |  |
| STK11 Mutation |  |  |  |
| Present | 0 (0.0%) | 0 (0.0%) | 1 |
| Absent | 15 (100.0%) | 5 (100.0%) |  |

| RHEB Mutation |  |  |  |
| --- | --- | --- | --- |
| Present | 0 (0.0%) | 0 (0.0%) | 1 |
| Absent | 15 (100.0%) | 5 (100.0%) |  |
| RICTOR Mutation |  |  |  |
| Present | 0 (0.0%) | 0 (0.0%) | 1 |
| Absent | 15 (100.0%) | 5 (100.0%) |  |
| MTOR Mutation |  |  |  |
| Present | 0 (0.0%) | 0 (0.0%) | 1 |
| Absent | 15 (100.0%) | 5 (100.0%) |  |
| RPTOR Mutation |  |  |  |
| Present | 1 (6.7%) | 0 (0.0%) | 1 |
| Absent | 14 (93.3%) | 5 (100.0%) |  |

**Table S6.** Comparison of Late-Onset PDAC Patients Treated with Gemcitabine Versus Those Not Treated with Gemcitabine.

| PI3K Pathway |  |  |  |
| --- | --- | --- | --- |
| Gene | Late-Onset<br>Treated with Gemcitabine<br>n (%) | Late-Onset<br>Not Treated with Gemcitabine<br>n (%) | p-value |
| PTEN Mutation |  |  |  |
| Present | 0 (0.0%) | 0 (0.0%) | 1 |
| Absent | 91 (100.0%) | 73 (100.0%) |  |
| PIK3R1 Mutation |  |  |  |
| Present | 1 (1.1%) | 0 (0.0%) | 1 |
| Absent | 90 (98.9%) | 73 (100.0%) |  |
| PIK3R2 Mutation |  |  |  |
| Present | 1 (1.1%) | 0 (0.0%) | 1 |
| Absent | 90 (98.9%) | 73 (100.0%) |  |
| PIK3R3 Mutation |  |  |  |
| Present | 1 (1.1%) | 0 (0.0%) | 1 |
| Absent | 90 (98.9%) | 73 (100.0%) |  |
| PIK3CA Mutation |  |  |  |
| Present | 4 (4.4%) | 0 (0.0%) | 0.1295 |
| Absent | 87 (95.6%) | 73 (100.0%) |  |
| INPP4B Mutation |  |  |  |
| Present | 1 (1.1%) | 0 (0.0%) | 1 |
| Absent | 90 (98.9%) | 73 (100.0%) |  |
| AKT1 Mutation |  |  |  |
| Present | 0 (0.0%) | 0 (0.0%) | 1 |
| Absent | 91 (100.0%) | 73 (100.0%) |  |
| AKT2 Mutation |  |  |  |
| Present | 1 (1.1%) | 0 (0.0%) | 1 |
| Absent | 90 (98.9%) | 73 (100.0%) |  |
| AKT3 Mutation |  |  |  |
| Present | 1 (1.1%) | 0 (0.0%) | 1 |
| Absent | 90 (98.9%) | 73 (100.0%) |  |
| PPP2R1A Mutation |  |  |  |
| Present | 2 (2.2%) | 0 (0.0%) | 0.503 |
| Absent | 89 (97.8%) | 73 (100.0%) |  |
| TSC1 Mutation |  |  |  |
| Present | 1 (1.1%) | 0 (0.0%) | 1 |
| Absent | 90 (98.9%) | 73 (100.0%) |  |
| TSC2 Mutation |  |  |  |
| Present | 2 (2.2%) | 0 (0.0%) | 0.503 |
| Absent | 89 (97.8%) | 73 (100.0%) |  |
| STK11 Mutation |  |  |  |
| Present | 2 (2.2%) | 2 (2.7%) | 1 |
| Absent | 89 (97.8%) | 71 (97.3%) |  |
| RHEB Mutation |  |  |  |
| Present | 0 (0.0%) | 0 (0.0%) | 1 |
| Absent | 91 (100.0%) | 73 (100.0%) |  |
| RICTOR Mutation |  |  |  |
| Present | 2 (2.2%) | 0 (0.0%) | 0.503 |
| Absent | 89 (97.8%) | 73 (100.0%) |  |
| MTOR Mutation |  |  |  |
| Present | 3 (3.3%) | 0 (0.0%) | 0.2545 |
| Absent | 88 (96.7%) | 73 (100.0%) |  |
| RPTOR Mutation |  |  |  |
| Present | 1 (1.1%) | 0 (0.0%) | 1 |
| Absent | 90 (98.9%) | 73 (100.0%) |  |

**Table S7.** Comparison of Early-Onset PDAC Patients Versus Late-Onset PDAC Patients Treated with Gemcitabine.

| PI3K Pathway |  |  |  |
| --- | --- | --- | --- |
| Gene | Early-Onset<br>Treated with Gemcitabine<br>n (%) | Late-Onset<br>Treated with Gemcitabine<br>n (%) | p-value |
| PTEN Mutation |  |  |  |
| Present | 0 (0.0%) | 0 (0.0%) | 1 |
| Absent | 15 (100.0%) | 91 (100.0%) |  |
| PIK3R1 Mutation |  |  |  |
| Present | 0 (0.0%) | 1 (1.1%) | 1 |
| Absent | 15 (100.0%) | 90 (98.9%) |  |
| PIK3R2 Mutation |  |  |  |
| Present | 0 (0.0%) | 1 (1.1%) | 1 |
| Absent | 15 (100.0%) | 90 (98.9%) |  |
| PIK3R3 Mutation |  |  |  |
| Present | 0 (0.0%) | 1 (1.1%) | 1 |
| Absent | 15 (100.0%) | 90 (98.9%) |  |
| PIK3CA Mutation |  |  |  |
| Present | 1 (6.7%) | 4 (4.4%) | 0.5411 |
| Absent | 14 (93.3%) | 87 (95.6%) |  |
| INPP4B Mutation |  |  |  |
| Present | 0 (0.0%) | 1 (1.1%) | 1 |
| Absent | 15 (100.0%) | 90 (98.9%) |  |
| AKT1 Mutation |  |  |  |
| Present | 0 (0.0%) | 0 (0.0%) | 1 |
| Absent | 15 (100.0%) | 91 (100.0%) |  |
| AKT2 Mutation |  |  |  |
| Present | 0 (0.0%) | 1 (1.1%) | 1 |
| Absent | 15 (100.0%) | 90 (98.9%) |  |
| AKT3 Mutation |  |  |  |
| Present | 0 (0.0%) | 1 (1.1%) | 1 |
| Absent | 15 (100.0%) | 90 (98.9%) |  |
| PPP2R1A Mutation |  |  |  |
| Present | 0 (0.0%) | 2 (2.2%) | 1 |
| Absent | 15 (100.0%) | 89 (97.8%) |  |
| TSC1 Mutation |  |  |  |
| Present | 0 (0.0%) | 1 (1.1%) | 1 |
| Absent | 15 (100.0%) | 90 (98.9%) |  |
| TSC2 Mutation |  |  |  |
| Present | 0 (0.0%) | 2 (2.2%) | 1 |
| Absent | 15 (100.0%) | 89 (97.8%) |  |
| STK11 Mutation |  |  |  |
| Present | 0 (0.0%) | 2 (2.2%) | 1 |
| Absent | 15 (100.0%) | 89 (97.8%) |  |
| RHEB Mutation |  |  |  |
| Present | 0 (0.0%) | 0 (0.0%) | 1 |
| Absent | 15 (100.0%) | 91 (100.0%) |  |
| RICTOR Mutation |  |  |  |
| Present | 0 (0.0%) | 2 (2.2%) | 1 |
| Absent | 15 (100.0%) | 89 (97.8%) |  |
| MTOR Mutation |  |  |  |
| Present | 0 (0.0%) | 3 (3.3%) | 1 |
| Absent | 15 (100.0%) | 88 (96.7%) |  |
| RPTOR Mutation |  |  |  |
| Present | 1 (6.7%) | 1 (1.1%) | 0.2642 |
| Absent | 14 (93.3%) | 90 (98.9%) |  |

**Table S8.** Comparison of Early-Onset PDAC Patients Versus Late-Onset PDAC Patients Not Treated with Gemcitabine.

| PI3K Pathway |  |  |  |
| --- | --- | --- | --- |
| Gene | Early-Onset<br>Not Treated with Gemcitabine<br>n (%) | Late-Onset<br>Not Treated with Gemcitabine<br>n (%) | p-value |
| **PTEN Mutation** |  |  |  |
| Present | 0 (0.0%) | 0 (0.0%) | 1 |
| Absent | 5 (100.0%) | 73 (100.0%) |  |
| PIK3R1 Mutation |  |  |  |
| Present | 0 (0.0%) | 0 (0.0%) | 1 |
| Absent | 5 (100.0%) | 73 (100.0%) |  |
| PIK3R2 Mutation |  |  |  |
| Present | 0 (0.0%) | 0 (0.0%) | 1 |
| Absent | 5 (100.0%) | 73 (100.0%) |  |
| PIK3R3 Mutation |  |  |  |
| Present | 0 (0.0%) | 0 (0.0%) | 1 |
| Absent | 5 (100.0%) | 73 (100.0%) |  |
| PIK3CA Mutation |  |  |  |
| Present | 0 (0.0%) | 0 (0.0%) | 1 |
| Absent | 5 (100.0%) | 73 (100.0%) |  |
| INPP4B Mutation |  |  |  |
| Present | 0 (0.0%) | 0 (0.0%) | 1 |
| Absent | 5 (100.0%) | 73 (100.0%) |  |
| AKT1 Mutation |  |  |  |
| Present | 0 (0.0%) | 0 (0.0%) | 1 |
| Absent | 5 (100.0%) | 73 (100.0%) |  |
| AKT2 Mutation |  |  |  |
| Present | 0 (0.0%) | 0 (0.0%) | 1 |
| Absent | 5 (100.0%) | 73 (100.0%) |  |
| AKT3 Mutation |  |  |  |
| Present | 0 (0.0%) | 0 (0.0%) | 1 |
| Absent | 5 (100.0%) | 73 (100.0%) |  |
| PPP2R1A Mutation |  |  |  |
| Present | 0 (0.0%) | 0 (0.0%) | 1 |
| Absent | 5 (100.0%) | 73 (100.0%) |  |
| TSC1 Mutation |  |  |  |
| Present | 0 (0.0%) | 0 (0.0%) | 1 |
| Absent | 5 (100.0%) | 73 (100.0%) |  |
| TSC2 Mutation |  |  |  |
| Present | 0 (0.0%) | 0 (0.0%) | 1 |
| Absent | 5 (100.0%) | 73 (100.0%) |  |
| STK11 Mutation |  |  |  |
| Present | 0 (0.0%) | 2 (2.7%) | 1 |
| Absent | 5 (100.0%) | 71 (97.3%) |  |
| RHEB Mutation |  |  |  |
| Present | 0 (0.0%) | 0 (0.0%) | 1 |
| Absent | 5 (100.0%) | 73 (100.0%) |  |
| RICTOR Mutation |  |  |  |
| Present | 0 (0.0%) | 0 (0.0%) | 1 |
| Absent | 5 (100.0%) | 73 (100.0%) |  |
| MTOR Mutation |  |  |  |
| Present | 0 (0.0%) | 0 (0.0%) | 1 |
| Absent | 5 (100.0%) | 73 (100.0%) |  |
| RPTOR Mutation |  |  |  |
| Present | 0 (0.0%) | 0 (0.0%) | 1 |
| Absent | 5 (100.0%) | 73 (100.0%) |  |

**Figure S1.**
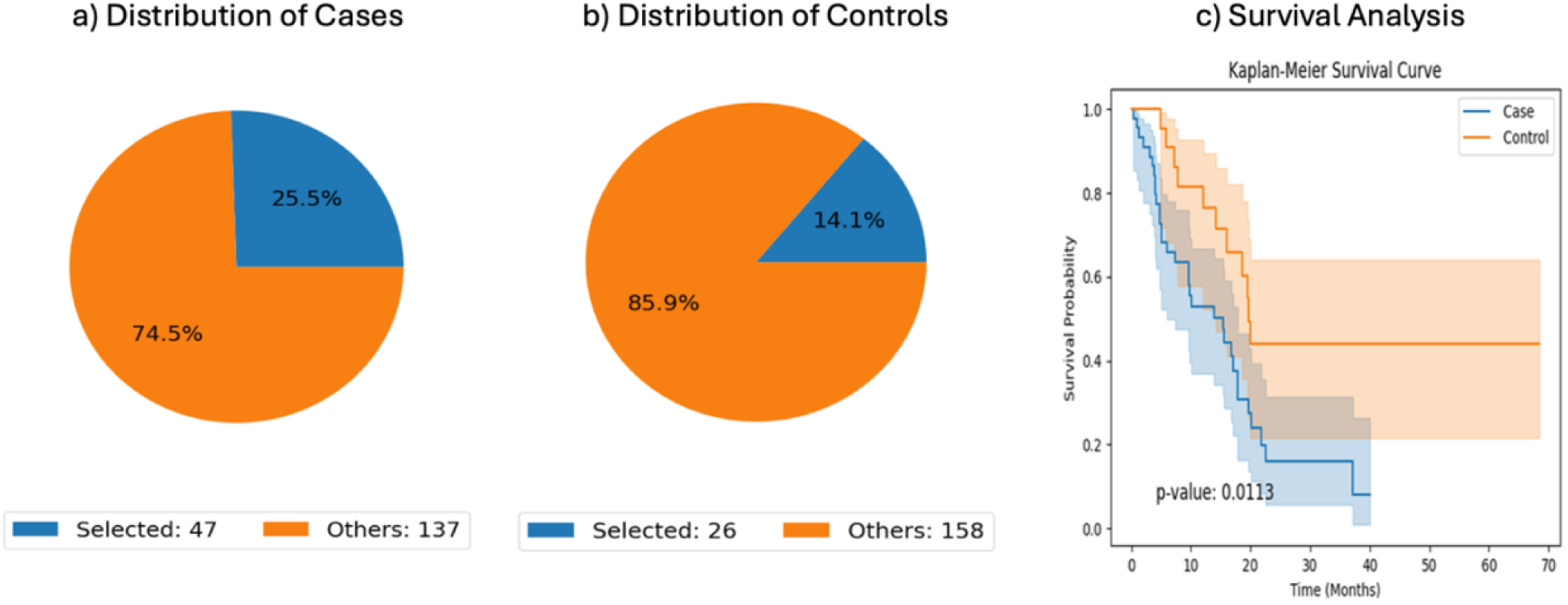
AI-guided cohort selection and overall survival comparison in late-onset PDAC not exposed to gemcitabine, stratified by TP53 pathway status. This supplementary figure shows how the conversational AI framework was used to define clinically specific cohorts and perform survival analysis in late-onset pancreatic ductal adenocarcinoma (PDAC) patients who did not receive gemcitabine. Based on natural language cohort criteria, the AI-HOPE-TP53 agent identified (a) a case group consisting of late-onset, non-gemcitabine-treated tumors with TP53 pathway alterations (n = 47; 25.5% of the full dataset) and (b) a control group composed of patients from the same clinical context whose tumors lacked TP53 pathway alterations (n = 26; 14.1% of the dataset). The pie charts summarize the proportion of selected and non-selected samples relative to the entire cohort. (c) Kaplan-Meier analysis demonstrated a significant difference in overall survival between these groups (log-rank p = 0.0113), with TP53 pathway-altered tumors associated with inferior survival compared with TP53 pathway-unaltered tumors. Shaded regions indicate 95% confidence intervals. These results support the prognostic relevance of TP53 pathway status in late-onset PDAC outside the setting of gemcitabine treatment and illustrate the ability of conversational AI to reproducibly generate clinically meaningful comparison cohorts.

**Figure S2.**
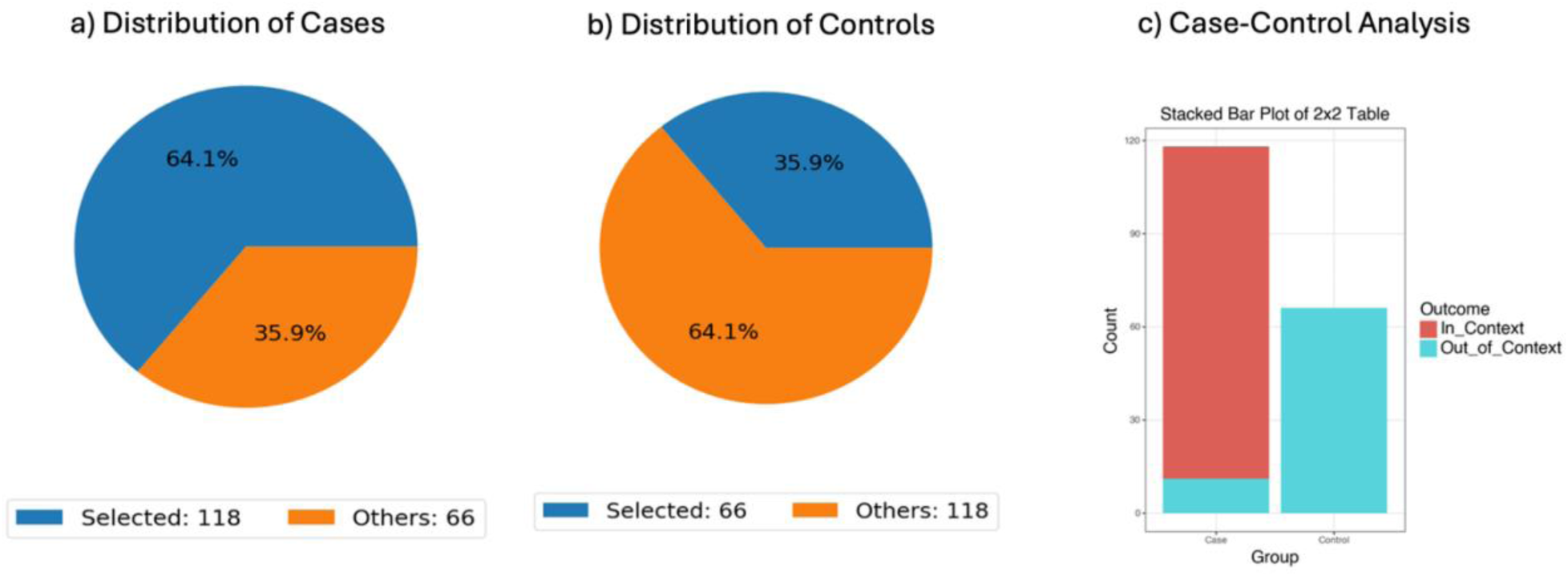
Conversational AI-driven comparison of TP53 pathway status within TP53-mutant PDAC tumors. This figure presents an AI-enabled enrichment analysis examining whether additional TP53 pathway alterations are differentially represented among pancreatic cancer patients whose tumors already harbor TP53 mutations. Using structured natural language criteria, the AI-HOPE-TP53 framework defined (a) a case cohort of TP53-mutant tumors with concurrent TP53 pathway alterations (n = 118; 64.1% of the dataset) and (b) a control cohort of TP53-mutant tumors lacking broader pathway-level alteration (n = 66; 35.9%). Pie charts illustrate the proportional distribution of selected versus non-selected samples across the full cohort. (c) A stacked bar plot summarizes the distribution of “in-context” (TP53 pathway-altered) and “out-of-context” (non-altered) samples across case and control groups. The majority of TP53-mutant tumors fall within the pathway-altered category, indicating substantial overlap between TP53 mutation and pathway-level dysregulation. Statistical comparison using Fisher’s exact test demonstrates a highly significant association (p ≈ 0), with an elevated odds ratio reflecting strong enrichment of TP53 pathway alterations among TP53-mutant cases. These findings reinforce the central role of TP53 as the dominant driver within its pathway and highlight how conversational AI can efficiently quantify relationships between gene-level mutations and broader pathway activation states in PDAC.

**Figure S3.**
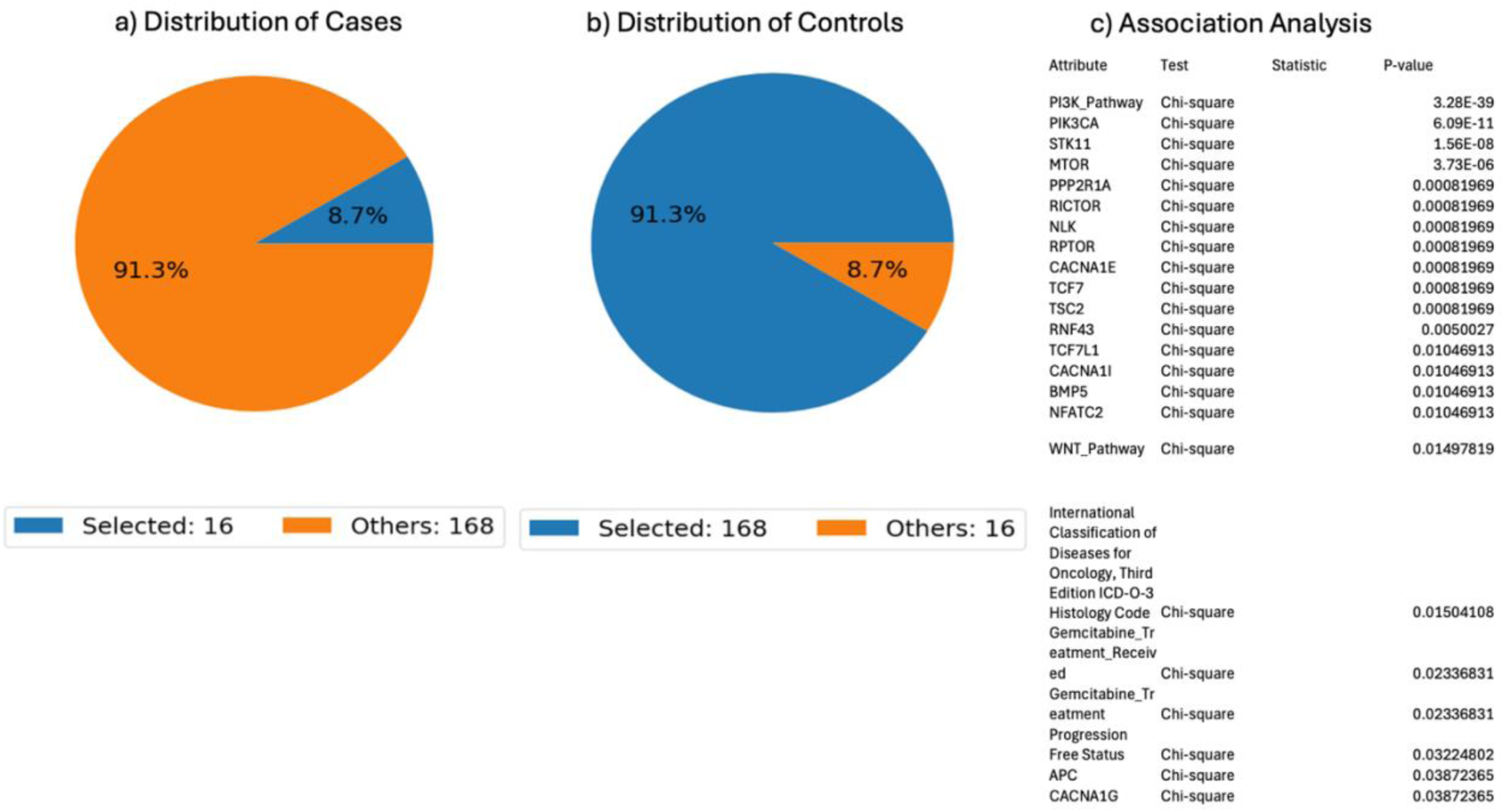
Conversational AI-based identification of clinical and molecular features associated with PI3K pathway alterations in PDAC. This figure summarizes an AI-guided comparative analysis of pancreatic ductal adenocarcinoma (PDAC) tumors stratified by PI3K pathway status across the full cohort. The case cohort comprised tumors with PI3K pathway alterations (n = 16; 8.7%), while the control cohort included tumors without detectable PI3K pathway alterations (n = 168; 91.3%). Panels (a) and (b) illustrate the relative proportion of selected and non-selected samples within each group, highlighting the comparatively low prevalence of PI3K pathway-altered tumors in this dataset. Panel (c) presents the results of a comprehensive association analysis using Chi-square testing to evaluate relationships between PI3K pathway status and a wide range of clinical and genomic attributes. Significant associations were observed with multiple PI3K pathway components and related signaling genes, including PIK3CA, STK11, MTOR, PPP2R1A, RICTOR, TSC2, and RPTOR, reflecting coordinated involvement of upstream regulators and mTOR complex-associated elements. Additional associations were identified with signaling and regulatory genes such as NLK, RNF43, TCF7, and members of the CACNA family, as well as broader pathway-level features including WNT pathway alterations. Clinically, PI3K pathway status was associated with treatment-related variables, including gemcitabine exposure, and with progression-free status, suggesting a potential link between PI3K pathway activation and disease behavior. Histologic classification (ICD-O-3) also demonstrated a significant association, indicating possible variation in PI3K pathway involvement across tumor subtypes. Overall, these findings indicate that PI3K pathway-altered PDAC represents a distinct, though relatively infrequent, molecular subset characterized by coordinated alterations across multiple signaling nodes and associations with treatment and clinical outcomes. This analysis highlights the capability of conversational artificial intelligence to efficiently uncover multidimensional relationships between pathway alterations and clinicogenomic features in complex cancer datasets.

**Figure S4.**
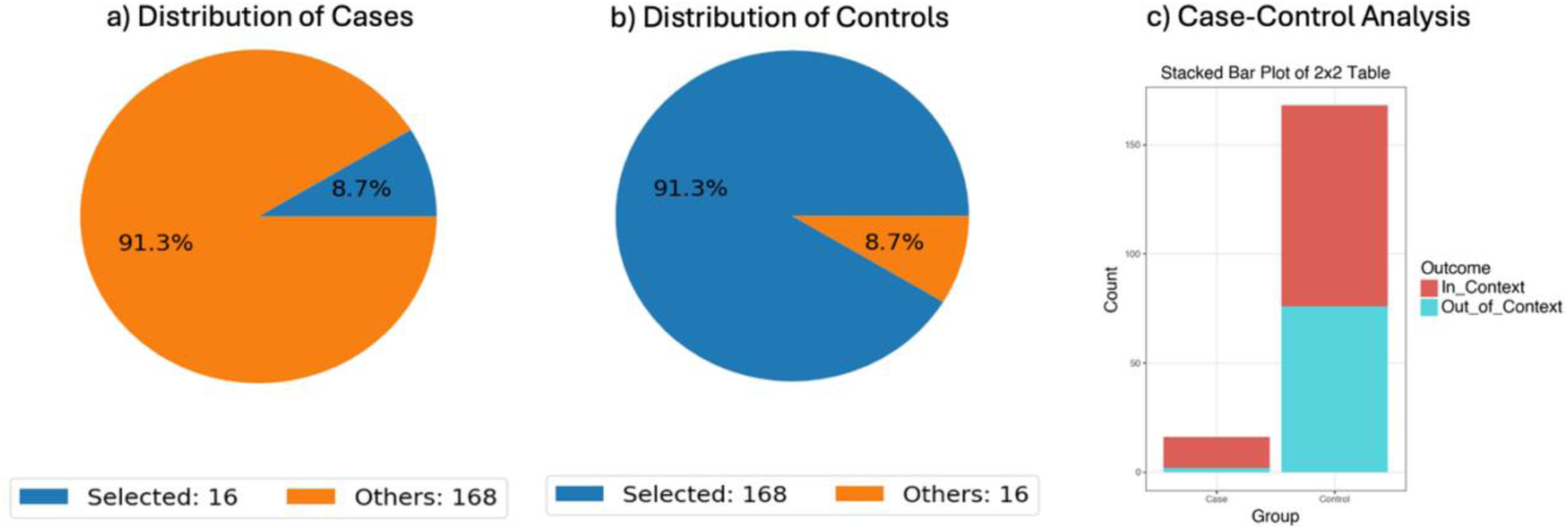
Conversational AI-based evaluation of gemcitabine exposure among PI3K pathway, defined PDAC subgroups. This figure illustrates an AI-driven odds ratio analysis assessing whether gemcitabine treatment is differentially represented between tumors with and without PI3K pathway alterations. Using structured query criteria within the AI-HOPE-PI3K framework, the case cohort included PI3K pathway-altered tumors (n = 16), while the control cohort consisted of PI3K pathway-unaltered tumors (n = 168). Pie charts in panels (a) and (b) depict the proportion of selected samples relative to the full dataset, highlighting the relatively small fraction of PI3K-altered cases. Panel (c) presents a stacked bar visualization comparing “in-context” (gemcitabine-treated) and “out-of-context” (non-treated) samples across both groups. Among PI3K-altered tumors, 87.5% were associated with gemcitabine exposure, compared with 54.76% in the PI3K-wild-type group. Statistical analysis demonstrated a significant difference between cohorts (Chi-square p = 0.023), with an odds ratio of 5.783 (95% CI: 1.274-26.24), indicating that PI3K pathway alterations are significantly enriched among gemcitabine-treated patients. These findings suggest a potential association between PI3K pathway activation and treatment exposure in PDAC, supporting the hypothesis that PI3K signaling may be linked to therapy-related tumor biology. More broadly, this analysis highlights the utility of conversational artificial intelligence for rapidly identifying treatment-associated molecular patterns within clinically stratified cancer datasets.

